# Mobility network reveals the impact of spatial vaccination heterogeneity on COVID-19

**DOI:** 10.1101/2021.10.26.21265488

**Authors:** Yuan Yuan, Eaman Jahani, Shengjia Zhao, Yong-Yeol Ahn, Alex Pentland

## Abstract

Mass vaccination is one of the most effective epidemic control measures. Because one’s vaccination decision is shaped by social processes, the pattern of vaccine uptake tends to show strong social and spatial heterogeneity, such as urban-rural divide and clustering. Examining through network perspectives, we develop a framework for estimating the impact of spatial vaccination heterogeneity on epidemic outbreaks. Leveraging fine-grained mobility data and computational models, we investigate two network effects—the “hub effect” (vaccinating mobility hubs reduces transmission) and the “homophily effect” (stronger homophily in vaccination rates increases transmission). Applying Bayesian deep learning and fine-grained epidemic simulations, our study suggests a negative effect of homophily and a positive effect of highly vaccinated hubs on reducing case counts for both the synthetic network and the U.S. mobility network. Our framework enables us to evaluate outcomes from various hypothetical spatial vaccine distributions and to study a hypothetical vaccination campaign strategy that targets a small number of regions with the largest gain in protective power using the data from January 2022. Our simulation suggests that our strategy can potentially prevent about 2.5 times more cases than a uniform strategy with an additional 1% of the population vaccinated. Notably, our simulation also shows that this strategy could even better protect vulnerable or disadvantaged communities through network effects than strategies that directly target them. Our study suggests that we need to examine the interplay between vaccination patterns and mobility networks beyond the overall vaccination rate, and that understanding geographical pattern of vaccine uptake could be just as important as improving the overall vaccination rate.

## Introduction

COVID-19 pandemic is not only a public health challenge but also an immense societal problem because social processes, such as political polarization and social contagion, significantly affect the course of epidemics^1–10^. Although the mass vaccination presents an effective road to the herd immunity ^11^, it is still challenging to predict the course of the pandemic as many sociopolitical factors come into play and multiple variants emerge^12–15^. These factors include highly unequal vaccine allocation across locations^16^, heterogeneous vaccine acceptance across social groups^14^, and their mixing patterns^17,18^ in social and mobility networks. Such heterogeneity raises important questions: What are the implications of heterogeneous spatial vaccine uptake across the society? How can the COVID-19 data inform us on improving vaccination campaign strategies for both the current and future pandemics?

Our study addresses these questions through large-scale epidemic simulations on the U.S. mobility network, using the observed and hypothetical vaccination distributions. Departing from highly aggregated models to understand vaccine performance^19–21^, we employ a data-driven approach to study the impact of spatial vaccination heterogeneity. Specifically, we leverage fine-grained human mobility data, vaccination data, and census data in the U.S. These rich datasets, along with fine-grained data-driven models^22–24^, enable us to study the outcome of hypothetical vaccination distributions and vaccination campaigns with unprecedented detail^14,18,25–27^.

The goal of our study is twofold—to develop a general network-based framework for analyzing the impact of spatial vaccination heterogeneity on transmission and to investigate the potential of location-based vaccination campaigns. Although our study uses the state of vaccination at a specific point in time, we show that our main conclusions are robust under different transmission dynamics and vaccination distributions, which suggest that and our methodology can be useful for future epidemics.

We begin by investigating the impact of spatial vaccination heterogeneity on COVID-19 by focusing on two major network effects. The first network effect is *homophily*, a phenomenon on the clustering of similar people, either due to sorting, social contagion, or local regulations^3,17,28^. In our context, homophily captures the fact that vaccination rates are similar among geographically close or socially connected locations^9,28,29^. A high level of homophily in vaccination leads to clusters of the unvaccinated, which may trigger localized outbreaks and produce more cases than expected by the overall vaccination rate. The second network effect is the *hub effect*, where the vaccination rate of central and highly mobile places can have a disproportionate impact on the case count^30,31^. Due to various reasons such as the urban-rural divide, hubs in the U.S. generally have a higher vaccination rate^32–34^, which may potentially reduce the severity of outbreaks. We visualize these two patterns at the county level in Figure 1 for illustration.

**Figure 1.**
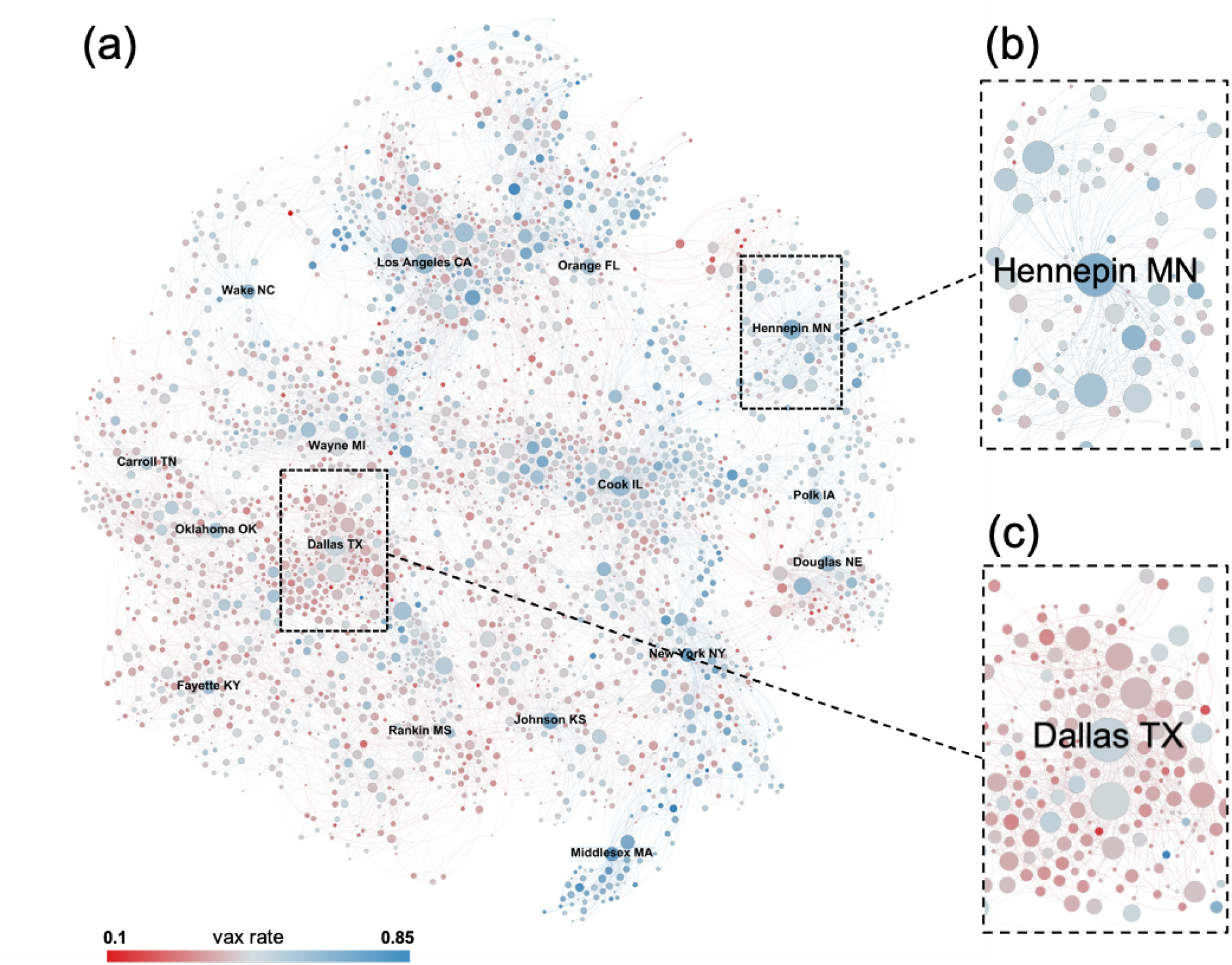
Illustration of the average vaccination rate in each county and the county-level mobility network backbone (a) among all U.S. counties, (b) for Hennepin county in MN and its adjacent counties, and (c) for Dallas county in TX and its adjacent counties. (see *Materials and Methods* for details). Nodes correspond to counties and are colored according to their vaccination rate, ranging from red (low) to blue (high), and are positioned according to the Fruchterman-Reingold layout^37^. The node size reflects its connectivity to other nodes. Panel (a) illustrates strong homophily, shown as localized clusters of the blue and red. For example, we see “blue clusters” for counties close to New York county in NY and Middlesex county in MA, while we observe “red clusters” for counties close to Dallas county in TX and Fayette county in KY. Panels (b) and (c) are the local networks for Hennepin county in MN and Dallas county in TX, respectively, where we observe that these hub counties that are connected to many other counties tend to have a higher vaccination rate than their adjacent counties.

To quantify the impact of these two effects on case counts, we examine both synthetic and fine-grained U.S. mobility networks. We design synthetic networks that exhibit either hub effect or homophily to study how these two effects operate in isolation. By comparing the original vaccination distribution with hypothetical vaccination distributions where we remove or flip the direction of homophily or hub effects, we find that the homophily effect exacerbates the size of an outbreak, while the hub effect attenuates it. Next, we repeat the same procedure on the empirical mobility networks and the COVID-19 vaccination distribution in the U.S. Because vaccination data is only available at the county level, we leverage additional fine-grained census features and Bayesian deep learning^35,36^ to infer vaccination rates at the level of the census block groups (CBGs). We show that given the vaccination distribution of COVID-19 in January 2022, the observed homophily accounts for at least a 9.3% increase in new COVID-19 infections within 30 days in comparison with hypothetical scenarios without homophily, while the hub effect caused by urban-rural divide substantially reduces the cases.

In the second part of our study, we address the question: where do increased vaccination rates reduce the largest number of cases in controlling the pandemic? Inspired by our findings on the network effects, we propose an efficient algorithm to find the locations that can maximally reduce the case numbers given a fixed increase in the overall population vaccination rate. While it is computationally challenging to search over all possible vaccination strategies based on transmission simulations for 200,000 CBGs, our algorithm solves these challenges by using gradient-based optimization on a differentiable surrogate objective. We estimate that such a strategy can reduce the number of cases by 9.5% with only a 1% increase in overall vaccination rate—an approximately 2.5 times improvement compared to vaccinating random locations. We also compare it with other reasonable vaccination strategies that do not take full network information into account, showing that targeting hubs or the least vaccinated locations are less effective. These results suggest that accurate location-based targeting can be a highly effective strategy to substantially reduce the size of outbreaks, although there are numerous societal, political and ethical issues to be addressed.

In addition to simply reducing the total number of cases, we should also consider the implications for vulnerable or disadvantaged communities, such as groups that are likely to experience severe illness, or regions with low vaccination rates. We examine case counts in groups that are stratified by age, race, income, locations with different vaccination rates. Our simulation suggests that the proposed targeting strategy reduces more cases in all groups compared to alternative strategies by the virtue of suppressing the overall spreading; for example, according to our simulations, the hypothetical strategy we study can better protect the communities with lowest vaccination rates than the strategy that directly targets them.

## Results

### Simulation results from two synthetic networks

We begin by employing synthetic mobility networks to study the impact of hubs and homophily on case counts. The goal of this first set of simulations is to isolate the impact of each effect by selectively controlling specific features of synthetic networks. To study the impact of homophily, we construct a *clustered network* of census block groups (CBGs) as nodes, where closely connected CBGs have a similar level of vaccination rates and the network is polarized into high and low vaccination regions. Separately, to show the impact of the hub effect, we construct a *centralized network* with a positive correlation between the network degree of a CBG and its vaccination rate. We then mix these two networks to observe how these two effects jointly affect the outcome. See *Materials and Methods* for the detailed explanation of how these synthetic networks are constructed.

To measure the impact of homophily or hubs on the severity of outbreaks, we “redistribute” vaccination over CBGs in a network that *removes* or *flips* either the homophily or hub effect, but otherwise is almost identical to the original vaccination distribution including the overall vaccination rate. We can then compare the outcome under a hypothetical vaccination distribution and the original vaccination distributions. Differences in the COVID-19 cases would estimate the impact of homophily or hub effects. Throughout the article, we consider four hypothetical vaccination distributions: “reverse”, “exchange”, “shuffle”, and “order.” We describe the construction of the hypothetical vaccine distributions in detail as follows:

- *Original*: It uses the original vaccination rates at the CBG level from the inference procedure discussed above.
- *Reverse*: If the original vaccination rate of a CBG *c* is *v*_*c*_, we assign 1 − *v*_*c*_ to it in the hypothetical network, thus “reversing” the vaccination rates. If hubs have high vaccination rates in the original scenario, they will end up with low rates in this hypothetical distribution. The homophily effect is preserved because by reversing all vaccination rates, the network assortativity of vaccination rates^38^ remains the same. We adjust the vaccination rates for all CBGs to match the average in the original distribution (the same for all other hypothetical distributions). ^*a*^
- *Exchange*: We “exchange” the vaccination rates of CBGs with similar mobility centrality scores. First, we rank all CBGs by their mobility centrality scores (see *SI* for the definition). Then, we conduct pairwise matching and exchange the vaccination rates of CBG pairs that are closest in their mobility centrality score^*b*^. In this way, we maintain the correlation between the mobility centrality score and the vaccination rate, while shuffling the vaccination rate distribution such that the homophily effect is reduced. Since the mobility centrality score follows the long tailed distribution, we do not exchange the CBGs in the top 1% mobility centrality scores, to avoid significant changes in the overall distribution of vaccination rates.
- *Shuffle*: We also randomly “shuffle” the vaccination rates among CBGs. This hypothetical distribution maintains the average and variance of the vaccination rates while simultaneously eliminating the homophily and hub effect.
- *Order*: We re-”order” the vaccination rates. First, we rank the CBGs by their mobility centrality scores; then for a CBG with a higher mobility centrality score (see *SI* for the definition), we assign a higher vaccination rate from the original distribution. In this way, the hypothetical distribution imposes the maximum level of the hub effect.

The vaccination rates in all hypothetical distributions is adjusted such that the CBG population-weighted average vaccination rate is the same as the original distribution: this would make any comparison across the hypothetical distributions appropriate as the total number of vaccinated people remains the same as the observed value and only its geographic distribution varies. The impact of all hypothetical distributions on homophily and the hub effect are summarized in Table 1.

**Table 1.** Impacts of four hypothetical vaccination distributions on homophily and the hub effect compared to the original distribution. +, -, =, × represent “increase”, “decrease’, “mostly unchanged”, and “flipped” respectively. Homophily in the networks is largely reduced or removed with “exchange”, “shuffle’, and “order”, but mostly unchanged by “reverse”; the hub effect is flipped by “reverse” (i.e., high vaccination CBGs become low vaccination in the hypothetical vaccination distribution and vice versa), removed by “shuffle”, improved by “order”, and mostly unchanged by “exchange”. The vaccination rates in all hypothetical distributions is adjusted such that the CBG population-weighted average vaccination rate is the same as the original vaccination distribution.

| Hypothetical vaccination distribution | Homophily | Hub Effect |
| --- | --- | --- |
| Reverse | = | × |
| Exchange | — | = |
| Shuffle | — | — |
| Order | — | + |

Figure 2 shows the simulated case counts under these hypothetical vaccination distributions, using a pre-assigned proportion of initially infected people (see *Materials and Methods* for details). The left panel shows any hypothetical vaccination distribution that removes the homophily effect (“exchange” or “shuffle”) reduces cases; this confirms our conjecture that stronger homophily increases the number of cases. Since there is no hub effect in this network, “reverse” has the same result as “original”, and introducing the hub effect (“order”) would greatly reduce the cases. The middle panel presents the simulated case counts on the centralized network which has a strong hub effect but no homophily. The main observation is that “shuffle” and “reverse” increase cases because they either eliminate or reverse the direction of the hub effect. Furthermore, since the network already has a perfectly strong hub effect, the “order” does not further improve the outcome. This confirms our assumption that the hub effect attenuates the severity of outbreaks. Since this network does not exhibit homophily, “exchange” does not have any impact.

**Figure 2.**
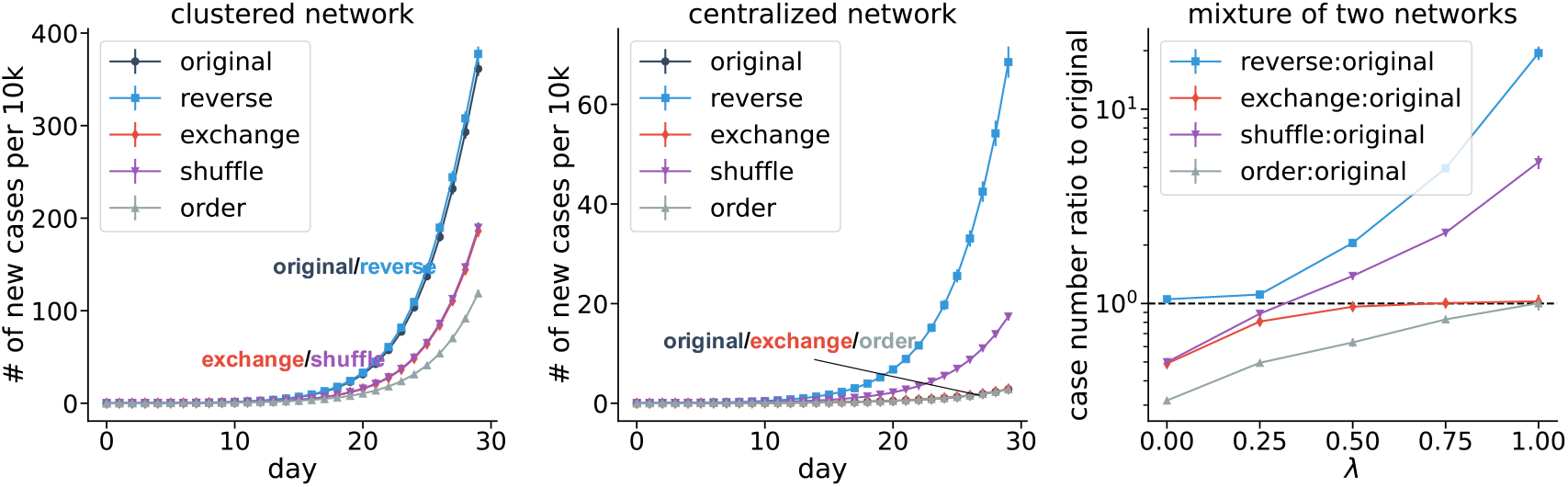
Simulation results for synthetic networks. (**Left**) For a network with strong homophily, both “exchange” and “shuffle” remove the homophily, leading to fewer cases. “Reverse” does not affect the homophily, as well as the outcome. “Order” slows down the spread by introducing the optimal hub effect. (**Middle**) For a network with strong hub effect, both “reverse” and “shuffle” remove the hub effect, leading to more cases. “Exchange” does not significantly affect the hub effect, nor the number of cases. (**Right**) A mixture of two networks (clustered and centralized networks) with a tuning parameter *λ* illustrates how the two effects can be mixed in a single network. Error bars are standard deviations from different runs of simulations.

The right panel shows how new infections change under the hypothetical vaccination distributions for different mixtures of the two synthetic networks. The network includes *λ* fraction of the edges from the centralized network and 1 − *λ* fraction of the edges from the clustered network and its vaccination rates are the weighted average of both networks (see *Materials and Methods* for details). As *λ* increases, the case count under the “reverse” hypothetical vaccination distribution increases due to a stronger hub effect. Similarly, when *λ* decreases, the case count under “exchange” decreases due to a stronger homophily. These results further verify that homophilous networks with vaccination clustering would have more cases whereas the highly vaccinated hubs decrease them.

The results in Fig. 2 allow for reinfection and breakthrough infection, as well as imperfect vaccine efficacy (70%), which reflect the attributes of COVID-19 as of January 2022. In *SI*, we show our conclusions remain robust when considering different vaccine efficacy levels or assuming no reinfection or breakthrough infection.

### Simulation results on the U.**S. mobility network**

Having illustrated the potential impacts of homophily and hubs, we then examine these effects on the observed U.S. mobility network and vaccination rates. The nationwide mobility network is constructed based on mobile phone users’ home census block group (CBG) and the points of interest (POIs) they visit on an hourly basis (see *Materials and Methods* for details). The goal of our study is to examine the impact of current vaccination patterns if human mobility were to recover the pre-pandemic levels (i.e. full reopening without masking or distancing requirements); thus we use the mobility data from 2019.

Since the vaccination rates are only available at the county level, we extrapolate them to the CBG level using additional CBG-level census demographic and spatial features. Our method uses a graphical model^39^ and Bayesian neural networks^35^ to capture the joint distribution between the observed variables (CBG-level census features and county-level vaccination rates) and the hidden variables (CBG-level vaccination rates), and infers the hidden CBG-level vaccination rates through variational inference^36,40^ (see *Materials and Methods* for the description of the algorithm). CBG-level vaccination rates are not publicly available so we cannot validate our predictions at the CBG level. Nevertheless, we validate the inferred vaccination rates from our method at the zip-code level (which is finer than county-level but coarser than CBG-level) by using available zip-code level data from a few states. Our method more accurately infers the zip-code level vaccination rate than alternative small area estimation methods based on linear or logistic regression (See *Materials and Methods*). Similar to the simulations on the synthetic network, we examine the impact of homophily and hub effects on case count using “reverse”, “exchange”, “shuffle”, and “order” hypothetical distributions described above. Here we apply the “reverse” hypothetical distribution to each state separately (see *Materials and Methods* for the rationale).

Figure 3 shows the simulated case counts over 30 days for the original and hypothetical scenarios (see *SI* for details of the simulation). Compared to the actual vaccine distribution, the “exchange” distribution reduces the cases by 9.3%. This result agrees with our conclusion from synthetic networks that homophily in vaccination rate adversely affects the number of new infections. In *SI*, we show that there is a high correlation between the vaccination rate of a CBG and that of its neighbors, implying a high level of homophily by vaccination, and the correlation largely disappears after the “exchange” indicating the successful removal of homophily in the network. The “reverse” distribution increases the cases by 13.2%, pointing to the positive impact of high vaccination rates in hubs compared with the original scenario. The “shuffle” distribution has a similar result with “original”, indicating the homophily effect may be of similar size with the hub effect. The “order” distribution demonstrates the huge potential of further exploiting the hub effect, since assigning the highest vaccination rates to central nodes reduces the case count by 30.4% compared to “original.” In *SI*, We also examine the predictions of our framework using the vaccination data from July 1st, 2021 and assuming perfect vaccines with no reinfection or breakthrough infection. The results from this alternative scenario remain the same, despite much stronger homophily and hub effects. In addition, we run separate simulations for each state to investigate how the homophily and hub effect affect the spread within each state, which offers insights into how our results might generalize to other regions or smaller countries.

**Figure 3.**
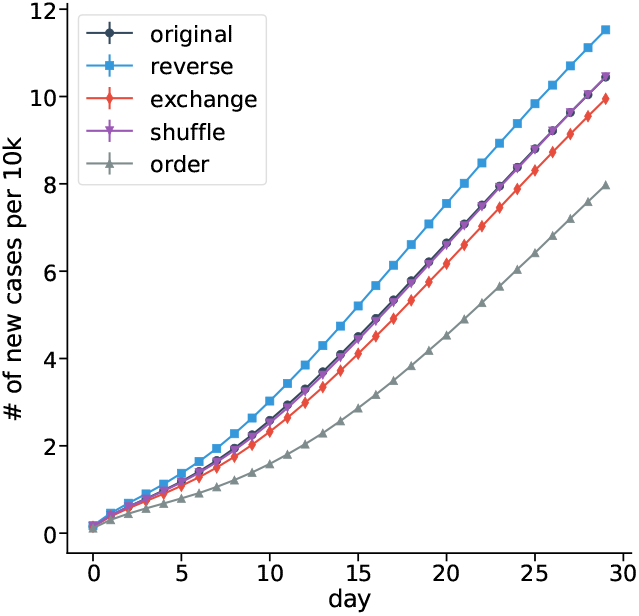
The number of simulated cases based on the U.S mobility network under various hypothetical scenarios. Error bars are standard deviations of simulations, which are in general smaller than the markers. These results support our hypothesis that the homophily effect harms the outcome while the within-state hub effect improves it. Specifically, by removing homophily, “exchange” reduces the cases. By flipping the hub effect, “reverse” increases the cases. The “order” hypothetical distribution strengthens the hub effect, and greatly reduces the number of cases; “shuffle” does not appear to change case counts.

### Informing effective vaccination campaigns

Motivated by the strong network effects found in previous sections, we explore the a hypothetical vaccination campaign strategy to maximally reduce the case counts by focusing on only a small number of CBGs. Such a vaccination campaign may target a specific set of census block groups that are well-connected or clustered with low vaccination rates. Here, our goal is to demonstrate the pivotal role that a small number of locations play in an epidemic, rather than providing concrete vaccination plans for the current pandemic. We acknowledge that any vaccination plan must consider numerous ethical issues (such as equitable vaccine distribution) before real-world implementation. Nevertheless, we report that the vaccination strategy we study here may better protect almost every demographic or geographic groups captured in our data when compared with the baseline strategies or existing vaccine roll-out strategies.

Deriving the case-optimized CBG targets is a significant computational challenge because it involves testing numerous combinations of thousands of CBGs out of over 200,000 CBGs in total. Therefore, our main technical contribution here is to design an algorithm that addresses this computational challenge by using the projected gradient descent^41,42^ to optimize a computationally feasible surrogate objective. To study the effectiveness of the targeted vaccination campaign, we compare it against four baseline policies: uniform targeting, random targeting, targeting least vaccinated CBGs, and targeting the most central CBGs. To guarantee a fair comparison, we assume that all policies induce the same number of people (an additional 1% of the U.S. population)^*c*^ to receive the vaccine. The details of our algorithm, the validation steps and the baseline policies are all presented in *Materials and Methods*. The simulation results given the vaccination state as of January 2022 are presented in the upper left panel of Figure 4. The uniform or the random vaccine campaign show the poorest performance among all strategies (2.7% reduction in case counts). Targeting the least vaccinated CBGs achieves slightly better performance, with a 3.4% reduction. Surprisingly, our proposed policy reduces 9.5% of cases, which is three times more than the uniform targeting or random strategies. Our proposed campaign also has a significantly better outcome than targeting the most central CBGs, which reduces cases by 8.1%.

**Figure 4.**
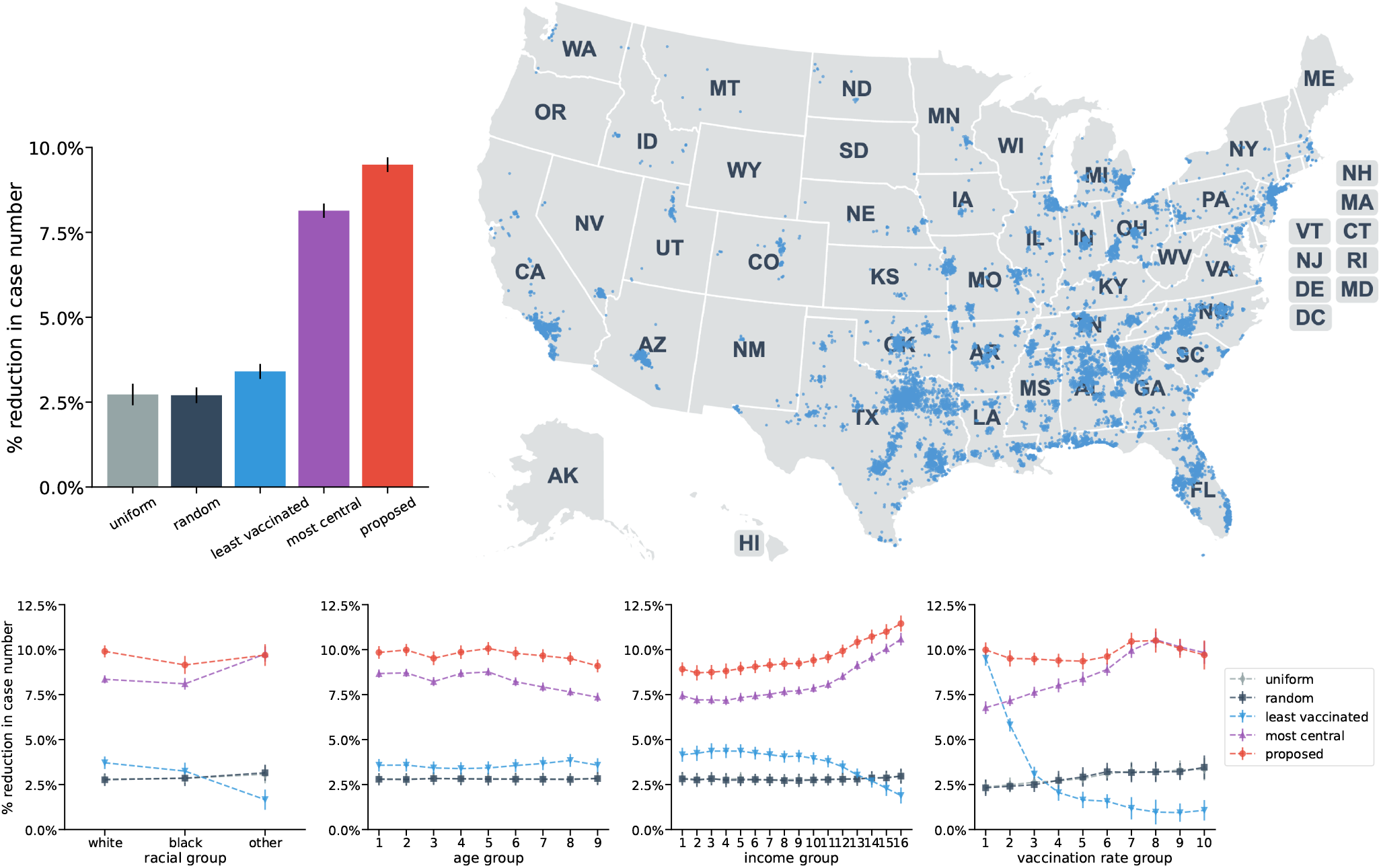
(**Upper left**) The performance of the five strategies given the state of January 2022. The *y*-axis is the reduction compared to the current vaccination distribution (“original”) in the first 30 days. Error bars are standard deviations from different runs of simulations. (**Upper right**) The most strongly targeted CBGs under our proposed strategy. Each blue dot represents a targeted CBG whose vaccination rate is proposed to increase by more than 5%. (**Lower**) Comparison of the five strategies across different demographic or geographic groups. Categorization of age and income groups are determined by the U.S. census. Vaccination rate groups of CBGs are determined based on equal-size deciles of the vaccination rate distribution, with higher groups corresponding to higher CBG vaccination rate.

While the hypothetical strategy we study is dependent on the ongoing pattern of spreading, surprisingly, we find that the targeted CBGs mostly remain the same under different transmission dynamics, reinfection rates, existing vaccination rates, as well as vaccination efficacy. Specifically, 87% of the targeted CBGs given the vaccination state as of January 2022 overlap with those obtained as of July 2021. Therefore, we expect that the same vaccination campaign strategy can be effective for different attributes of a pandemic.

An important question regarding the optimized vaccination campaign is whether it takes advantage of the hub effect or the homophily effect. The upper right panel of Figure 4 presents the map of the targeted CBGs. We observe that our policy primarily targets hub cities and the south (which includes several clusters with low vaccination). 80.1% of the targeted CBGs by the proposed policy overlap with those targeted by the most-central policy. This suggests that an ideal strategy mainly leverages the impact of the hub effect to improve the outcome of the vaccination campaign, echoing the “order” distribution result.

In addition to reducing case counts, a successful strategy should protect vulnerable populations and not exacerbate existing social inequalities. For example, it could be more important for a vaccination strategy to reduce cases among older population who are more vulnerable to severe illness or death. As another example, a vaccination campaign that only benefits high income groups is unacceptable. Because the case-optimized strategy we study here targets a very small number of places, especially the hub cities, it is critical to examine the impact of the strategy on a diverse array of sub-populations, especially disadvantaged populations. To further understand the strategy’s impact on equity, we present simulated case counts across demographic and geographic groups in the lower panel of Figure 4. We find that this hypothetical strategy Pareto-dominates baseline strategies, i.e., the case-optimized strategy reduces comparable or more case counts than baseline strategies on every demographic group that we could examine, by the virtue of substantially suppressing the epidemic. Moreover, compared with the strategy targeting the least vaccinated CBGs, this hypothetical strategy can protect the CBGs with the lowest vaccination rates even better. However, we should also note that this strategy, along with the strategy that targets the most central CBGs, tends to disproportionately benefit the high income groups. Although this is beyond the scope of this paper, this issue can potentially be addressed by modifying the objective function to account for vaccine equity.

## Discussion

While increasing the overall vaccination rate is crucial to the control of epidemics, results from our proposed framework show that the spatial heterogeneity of vaccination also has a strong impact on the course of the epidemic, and therefore should be understood well. Specifically, we show that, through both synthetically generated networks and real-world mobility networks, homophily can increase case counts whereas highly vaccinated hubs reduce them. Our framework can be easily updated to reflect new developments such as new variants as the size of these effects can vary for future waves of the pandemic. We also propose a promising method for identifying a small number of the most pivotal locations. We show that, when targeted, the increased vaccination in these locations has a disproportionate effect on suppressing the epidemic. These locations tend to be both the transportation hubs and the places that can also leverage the homophily effect.

Our study may be generalized beyond the U.S. and the current pandemic. In addition to the synthetic networks and the U.S. mobility network, we conduct the transmission simulation on the mobility network of each U.S. state, assuming no cross-state mobility, and observed similar results. Our main conclusions on network effects remain robust regardless of synthetic or the U.S. mobility networks, or consideration of vaccine efficacy, breakthrough or reinfection. These results show the potential generalizability of our main conclusions on network effects to other countries, future mobility trends, or other epidemics such as measles^43^.

Our results implies the existence of a substantial spatial heterogeneity regarding the impact of each additional vaccine. A small number of locations would have a disproportionate impact on the nationwide outcome. Thus, we argue that understanding the implications of the spatial distribution of vaccination is as important as that of the overall vaccination rate in predicting the course of the pandemic. We show that there may be a large, untapped potential to utilize the network effect and improve the effectiveness of the vaccination campaign. Even though it is sometimes suggested that one should focus on the least vaccinated places (in fact, current campaigns are already using this strategy^44^), our results suggest that this may not be the most effective strategy in reducing the total impact as well as helping the most vulnerable populations.

At the same time, our conclusions should be interpreted with caution. First, our mobility networks are constructed from data provided by SafeGraph, which collects human mobility data from mobile applications. The data may over-represent certain demographic groups who use mobile phones more frequently and are shown to miss records of non-POIs where infectious diseases also spread. Moreover, the quality of vaccination data and the predictability of simulation models may also affect our estimations. Therefore, we recommend using our results to theoretically demonstrate how network effects shape an epidemic, rather than quantitatively estimate the case counts directly used by policymakers. Furthermore, any real-world applications based on our study must examine their social implications and ethical concerns. For instance, the campaign strategy studied here may preferentially target certain socio-demographic groups and its benefits may not be equally distributed to different populations. It may be more fruitful to view our method as a way to identify locations that disproportionately affect the course of the epidemic. Future studies may explore alternative objective functions that take social equities or other quantities such as hospitalizations or deaths into account.

## Materials and Methods

### Data collection

The network is constructed using the U.S. mobility from SafeGraph, a company that provides aggregated data collected from mobile applications. All data is anonymized and aggregated by the company so that individual information is not re-identifiable. This dataset has been widely adopted to study human mobility patterns, particularly during the COVID pandemic ^1,3,24,45–48^. Most notably, an epidemic model built on this data—which we adopt in this paper—has shown to be highly predictive of the size of local outbreaks especially for the first wave of COVID-19, as well as other stylized facts^24^. SafeGraph receives the location data from “third-party data partners such as mobile application developers, through APIs and other delivery methods and aggregates them.” This data reflects the frequency of mobility between all points of interest (POIs) and the census block groups (CBGs) in the United States. Specifically, the data contains information on the number of people at a CBG who visit a POI on a certain day or in a certain hour. The data also contains the information for each CBG’s area, median dwell times, as well as geo-locations of all CBGs and POIs. In total, there are 214,697 CBGs and 4,310,261 POIs in the U.S. We use the 2019 mobility data to reflect the scenario when all businesses were to fully reopen and social distancing and masking are not imposed.

We also collected the latest U.S. census data from the SafeGraph database (the complete US Census and American Community Survey data from 2016 to 2019). The data contains the demographic features of each CBG, such as the fractions of each sex, age group, racial and ethnic group, education level, and income level. The vaccination data come from the Centers for Disease Control and Prevention (CDC)^*d*^ which provides daily vaccination records on all states except Hawaii ^*e*^. The results in the main text reflect the state of vaccine distribution from January 21st, 2022, with considerations of imperfect vaccine, reinfection, and breakthrough cases. In *SI*, we also examine the model predictions using the vaccination data from July 1st, 2021 assuming perfect vaccine efficacy and no reinfection; our conclusions from both of these simulations remain robust.

### Constructing mobility network of counties or CBGs

Our simulation setup follows ref. 24 where we first construct a *mobility bipartite network* for a given region (country or state),. The edges in the bipartite network are between POIs (denoted by the set *𝒫*) and CBGs (denoted by the set *𝒞*). The edge weight between a POI *p* ∈ *𝒫* and a CBG *c* ∈ *𝒞* corresponds to the number of people who live in CBG *c* and visit POI *p*. The bipartite network can vary over time according to the SafeGraph mobility data. However, since our study aims to illustrate the two network effects rather than to provide exact predictions in growth of COVID-19 cases, we aggregate the hourly number of visits in 2019 and construct the bipartite network for each hour given the annual average weights (persons per hour).

The undirected *mobility network among CBGs* is derived by projecting the aforementioned bipartite graph, considering the areas and dwell times of each POI. In this network, the edges between two CBGs *c* and *c*′ is

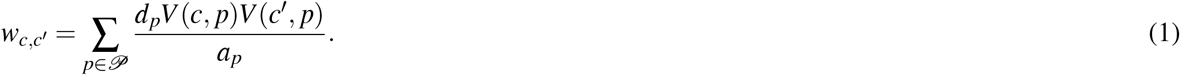

Here *p* corresponds to a POI, *V* (*c, p*) is the hourly average number of visitors from CBG *c* at POI *p, a*_*p*_ is the area of POI *p. d*_*p*_ is the probability of two people visiting the POI *p* at the same time, derived from the median dwell time at the POI as described in ref. 24. This edge weight is consistent with the simulation process proposed by ref. 24, as illustrated later in Eq. (2). The edge weight is proportional to the number of people in CBG *c* who get infected from CBG *c*′ assuming equal infection rate across all CBGs. The network in Figure 1 is constructed using the same method, but it is at the level of counties instead of CBGs for illustrative purposes. We only retain the top five neighbors of each county with the highest weights (thus making it as a directed graph); and then we convert the directed graph to an undirected one for Fig. 1. We use the full CBG-level network in our simulations.

### The synthetic networks

In this section, we explain the construction of the synthetic networks in detail. The construction of networks is consistent with the input in the model of ref. 24, where the basic element is a CBG and individuals are homogeneous among each CBG. For the clustered network, we first generate 10,000 CBGs, each with 10,000 residents. These CBGs are equally divided into 100 clusters (by cluster id), each of which can be considered as, for example, a “city.” Similarly, we create 10,000 POIs, which are equally spread out in the 100 clusters. All POIs have identical areas and dwell times. We next generate the edges. People living in one CBG visit the 100 POIs within the same cluster with a high probability (for each POI with an hourly probability of 40%), but visit the other 9,900 POIs with a small probability (for each POI with an hourly probability of 0.05%). We draw from the Bernoulli distribution to determine whether there exists at least one person from the CBG who visits the POI. To create some heterogeneity in the number of visits, the number of additional visitors follows the Poisson distribution Pois(1). To create the homophily effect, we randomly assign vaccination rates (either 80% or 20%) to clusters. That is, all CBGs in the same cluster have the same vaccination rate, which is either 80% and 20%, which creates a strong contrast of vaccination rates among different groups.

For the centralized network, we also assume there are 10,000 CBGs and 10,000 POIs, with a population of 10,000 in each CBG. However, instead of organizing into clusters of similar vaccination, the CBGs exhibit a high level of variation in their degree centrality. We first generate the random variable *D*_*c*_ for each CBG *c*, which follows a power distribution (with density function 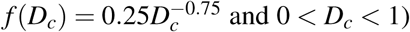). The CBG is then connected to 100 × *D*_*c*_ (rounded to an integer) randomly selected POIs. In this way, the degree distribution will be skewed with considerable degree heterogeneity among CBGs—a few CBGs will connect to many POIs, thus becoming central, while the majority will connect to only a few POIs. Similar to the clustered network, the number of people who visit each POI from CBG *c* follows the Poisson distribution Pois(1), which generates a certain level of heterogeneity in the edge weights. To create a positive hub effect, we impose a positive correlation between the degree of the CBG and its vaccination rate. In particular, the vaccination rate of a CBG *c* is set to be 0.4 + 0.5*D*_*c*_, thus ensuring that more central CBGs (with higher degree) have higher vaccination rates. Finally, since the degree of each CBG and in particular the set of POIs it is connected to is independent of other CBGs, the network will not exhibit homophily in vaccination.

Finally, hybrid networks (with varying *λ*) are constructed by mixing the clustered and centralized networks with a parameter *λ* that controls the composition of the mixture. We first randomly map each CBG in the instance of the clustered network to a CBG in the instance of the centralized network.^*f*^ For a given value of *λ*, an edge between a CBG and a POI in the clustered network is kept with a probability of *λ* independent of other edges, and similarly an edge from the centralized network is kept with probability of 1 − *λ*. Thus, a higher value of *λ* implies a stronger hub effect and a weaker homophily. The vaccination rate of a CBG is the weighted average of the vaccination rates in the centralized network and the clustered network, with weights of *λ* and 1 − *λ* respectively. As *λ* increases, the correlation between centrality and the vaccination rate increases and the degree of homophily decreases.

### Inferring CBG-level vaccination rate

County-level vaccination rates are provided by the CDC on a daily basis, while fine-grained CBG-level vaccination rates are unavailable. Because counties cover relatively large, heterogeneous areas and because the epidemic model we use is formulated at the level of CBGs, which offers a much higher resolution than county-level models and predicts the epidemic growth with high accuracy, we estimate the CBG-level vaccination rates from county-level vaccination rates.

This problem is called “small area estimation”^49^, where the goal is to use aggregated statistics (such as county-level vaccination rate) and socio-demographic characteristics to infer corresponding statistics at a more fine-grained resolution (such as CBG-level vaccination rate). To enable more accurate inferences, we use demographic and geographic features such as sex, age, race and ethnicity, income level, education level, and geographical coordinates, which are available for all the CBGs.^*g*^ Our assumption is that CBGs with similar features should have similar vaccination rates. This is a missing data imputation problem as illustrated in Fig. 5, where the observed variables are county-level vaccination rates and CBG-level features, while the missing variables are the CBG-level vaccination rates.

**Figure 5.**
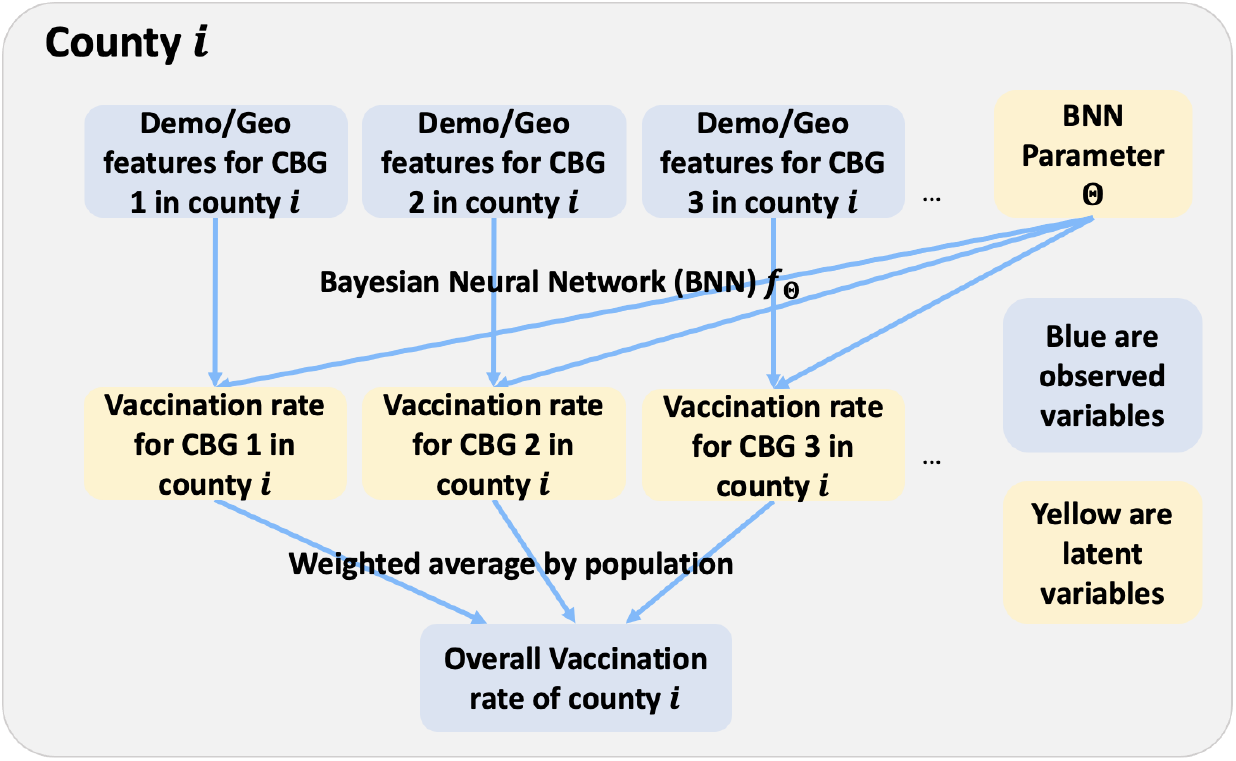
A Bayesian latent variable model to impute the CBG-level vaccination rate from the county-level vaccination rate. For each county (indexed by *i*) we observe the county-level average vaccination rate; for each CBG we observe demographic and geographic features (proportions of different sex, age, racial or ethnic, income, and education groups as well as the geo-locations). The latent variables (which we need to impute) are the vaccination rate for each CBG. We model the mapping from each CBG’s feature to vaccination rate as a Bayesian neural network with unknown parameters Θ. Given the observed variables (blue boxes) we infer the posterior distribution on the latent variables (yellow boxes).

We design a Bayesian model shown in Fig. 5 to impute the missing variables (i.e., the CBG-level vaccination rates). The benefit of the Bayesian approach is that once we define the data generation process, we can compute the Bayesian posterior over the missing variables given the observed variables with standard inference methods^36^. We define the following data generation process: for each CBG we observe the demographic and geographic features; the features are inputs to a Bayesian neural network^35^ with unknown parameter Θ, which outputs the vaccination rate of the CBG. Finally, we average the vaccination rates of all CBGs in a county to obtain the overall vaccination rate of that county. Since the posterior inference is approximate, the weighted average of CBG-level vaccination rates in a county does not exactly match the ground truth vaccination rate for that county. Thus, we rescale the inferred vaccination rates to match the ground truth county level vaccination rate. The algorithm is run for all CBGs in the U.S. simultaneously. Finally, we further improve performance slightly by ensembling multiple inferred vaccination rates from random initialized approximate inference procedures. In *SI* Fig. S8, we present examples of our inferred results. We use the interpolated CBG-level vaccination rates as the input for the downstream simulation tasks.

A major challenge is the performance evaluation because no CBG-level groundtruth data is available. We thus resort to validate on the zip code level groundtruth data. A county typically consists of multiple zip codes and a zip code corresponds to multiple CBGs. We aggregate predicted CBG-level vaccination rates to the predicted zip-code-level vaccination rate. Then we compare our predictions with the groundtruth on the zip code level. As of January 21st, 2022, the following states provide zip code level vaccination rates: California, Idaho, Illinois, Maine, New York, Oregon, Pennsylvania, and Texas. We thus only test on these states. Our approach has an MAE (weighted by zip code population) of 8.9%, which proves a 9.1%’s improvement over directly using the county level vaccination rates on the relative scale. In *SI*, we provide more details of this validation process and results.

### Simulating COVID-19 spreading

We extend the model in ref. 24 to simulate the spreading of COVID-19. The model is essentially an SEIR model^50^, but it is based on the full human mobility data at the level of CBGs and the key parameters in the SEIR model are estimated from the mobility network using machine learning tools. Susceptible individuals (S) first get exposed (E) to the disease with a certain probability after contacting infected people; then exposed people develop symptoms (I, infected) after a period of time; finally, the infected people get recovered or removed (R) after a period of time.

The key difference in our approach is that we also incorporate the vaccination status of individuals in the model using the CBG-level vaccination rate. For example, if a CBG *c* has a vaccination rate *v*_*c*_, we assume that a fraction (*αv*_*c*_) of individuals in the CBG are “recovered” at time 0. This implies that the vaccine efficacy is *α*. Note that because of the unavailability of more fine-grained data, the simulation does not consider any heterogeneity within a CBG. In other words, we assume that all individuals within a CBG have equal probability of getting vaccinated or infected.

The number of people in CBG *c* who newly get exposed (and then infected) at time *t* from POI *p* follows a Poisson distribution as shown below:

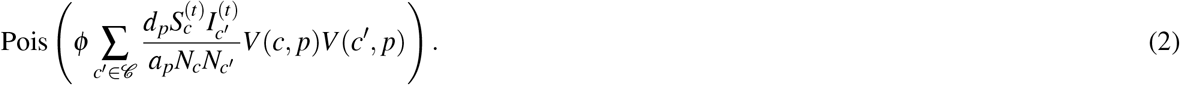

Here we follow the convention, using 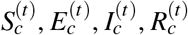 to denote the number of people in CBG *c* who are susceptible, exposed, infectious, and removed at the time stamp (i.e., hour) *t*, respectively. Other variables were defined along with Eq. (1). All exposed people will eventually become infectious, and all infectious will eventually become “recovered.”

Additionally, different from the original model in ref. 24, our study takes into account breakthrough infection and reinfection^51^. That is, recovered cases (naturally or vaccine-induced immune) may eventually return to being “susceptible.” In our model, the number of people in CBG *c* who switch from “recovered” to “susceptible” follows a Binomial distribution:

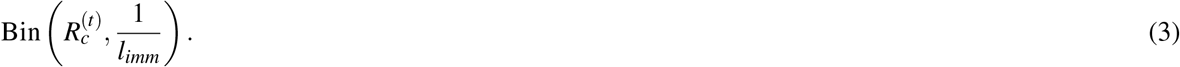

Here *l*_*imm*_ indicates the average length of immunity period after recovery or vaccination. For the parameter choices, please refer to *SI*.

Equation 2 also motivates our construction of edge weights for the network. The number of people in CBG *i* who get infected because of their contact with people from CBG *j* is proportional to the edge weight we define in Equation 1. This also inspired us to define the mobility centrality of a CBG (see *SI*).

### Designing the vaccination campaign

Here we explain the selection procedure of targeted CBGs in our proposed vaccination campaign strategy. Let *u* be the vector of the initial fraction of unvaccinated for each CBG (i.e., one minus the vaccination rate), and *v* be the increase in the vaccination rate under the campaign. Thus, *u* - *v* is the unvaccinated fraction vector after the campaign. Our goal is to find the optimal *v*^*^ that decreases the case count as much as possible.

The quantity (*u* - *v*)^*T*^*W* (*u* - *v*) is our objective function, which captures the growth rate of the cases. The intuition is as follows. First, from Eq. (2) we know that the number of people in CBG *c* who get infected from people in CBG *c*′ is proportional to 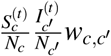. Under the “perfect” vaccination (i.e., vaccinated people do not get infected), we assume 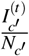 is highly correlated with (or approximately proportional to) the fraction of unvaccinated in *c*′, which is 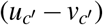; and 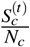 is highly correlated with (or approximately proportional to) the unvaccination rate of *c*, which is (*u*_*c*_ - *v*_*c*_). In other words, the unvaccination rate of a CBG predicts its factions of susceptible and infected population. Therefore, the value 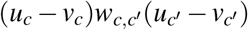 reflects the transmission from CBG *c* to *c*′. Using the matrix notation, (*u* - *v*)^*T*^*W* (*u* - *v*) is approximately proportional to the total transmission for all possible *c, c*′ pairs, or the number of new cases.

Intuitively, this objective function incorporates both the hub effect and homophily. For the hub effect, the increase in the vaccination rate of a CBG (by *v*_*c*_) reduces the objective function by *v*_*c*_ times the mobility centrality score of the CBG (see *SI*). Therefore, the optimization tends to improve the vaccination rates of more central CBGs. For the homophily effect, an increase in a CBG *c*’s vaccination rate results in the decrease of the objective function that is proportional to 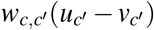 for all other *c*′ that are connected to *c*. Therefore, reducing the vaccination rate of one CBG spills over to the adjacent CBGs. The spillover effect is larger if the targeted CBG *c* is in a cluster of CBGs with similarly low vaccination rates. Thus, the optimization can exploit the homophily in the network by targeting clusters of low vaccination and further reducing the objective function by the spillover effect.

In addition, we impose several feasibility constraints. Specifically, we assume that *u* − *v* ≽ 0, which means that no CBG’s unvaccination rate is negative. Also, *v* ≽ 0, which indicates that vaccination campaign only reduces unvaccination rate and never increases it (i.e. always increases the vaccination rates). We also impose constraints that make the practical implementation of the vaccination campaign possible: specifically, it is difficult to decrease the unvaccination rate of a CBG by a large amount; a 10% increase in the vaccination rate of two CBGs might be much easier than a 20% increase in one CBG. Therefore, we require *v* ≼ 0.1, i.e., we reduce unvaccination rate of each CBG only up to 10%. Finally, to model finite resources, we limit the total number of vaccine doses to administer by *θ*, that is ⟨*v, m*⟩ ≤ *θ* where *m* is the population vector of CBGs. For our results, we set *θ* to 1% of the total population of the country, in other words, the proposed strategy increases the country-wide vaccination rate by at most 1%. Accordingly, the proposed strategy is the solution of the following optimization problem:

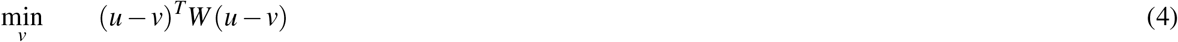

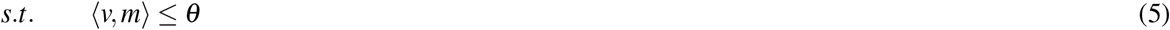

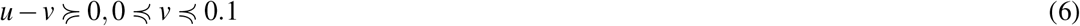

In *SI*, we provide more technical details of the optimization approach, and prove that we can find the global optimum of the optimization problem with projected gradient descent.

Finally, we compare the eventual case counts under the following five campaign policies by running the simulations under the same setting on the U.S. mobility data:

- *Proposed*. It uses the increase in vaccination rate of targeted CBGs proposed by our algorithm. The total number of targeted people is capped at 1%.
- *Uniform*. It increases the vaccination rates of all CBGs by 1%.
- *Random*. It increases the vaccination rate of randomly chosen CBGs by 10%. This process continues until an additional 1% of the whole population has been vaccinated.
- *Least vaccinated*. It increases the vaccination rate of CBGs with the lowest vaccination rate by 10%. The process of choosing the least vaccinated CBGs continues until an additional 1% of the whole population is targeted.
- *Most central*. It increases the vaccination rate of CBGs with the highest mobility centrality by 10%. The process of choosing the most central CBGs continues until an additional 1% of the whole population is targeted.

Note that the number of targeted CBGs may be slightly different across different policies, but the affected population remains the same.

## Data Availability

All data produced are available online at
https://github.com/yuany94/covid-vaccine

## Acknowledgements

We thank Ana Bento, Esteban Moro, and Christos Nicolaides for their helpful comments.

## Data availability

The U.S. Mobile phone mobility data and census data are available through the SafeGraph COVID-19 Data Consortium (https://www.safegraph.com/covid-19-data-consortium). Vaccination data are available on CDC website (https://covid.cdc.gov/covid-data-tracker/#datatracker-home). Synthetic data and the intermediate data generated from the aforementioned public is publicly available on GitHub: https://github.com/yuany94/covid-vaccine. All zip code level vaccination rates are retrieved from state health websites and are also available on GitHub.

## Code availability

Code is publicly available on GitHub: https://github.com/yuany94/covid-vaccine.

## Supplementary Information

### Replication of July 1st, 2021, without consideration of reinfection and vaccine efficacy issue

To demonstrate the robustness of our results, we also examine an earlier state of COVID-19: We collected the vaccine distribution data as of July 1st, 2021 and also applied our deep learning approach to estimate the CBG-level vaccination rates. Moreover, since we can safely assume that reinfection or breakthrough infection cases were uncommon given the state as of July 2021, our model does not take them into account for this July 2021 scenario (i.e., we no longer have Eq. (3 in the simulation). Moreover, for the state as of July 2021, we assume perfect vaccine – *α* = 1). Here we replicate the main text results for Figures 2 and 4 in Fig. S1 for the state as of July 1st, 2021.

Figure S1 shows that our main conclusions remain largely robust under different spatial vaccine distributions, variants, and vaccine efficacy levels. The upper left panel demonstrates the robustness of conclusions on hypothetical vaccine distributions. In particular, since we consider the vaccine efficacy perfect on July 1st, 2021, the discrepancy in the results of different hypothetical vaccine distributions appears to be even larger. Specifically, compared to the actual vaccine distribution, the “exchange” distribution reduces the cases by 22.0%. The “shuffle” distribution has a similar result with “original”, indicating the homophily effect may be of similar size with the hub effect., which simultaneously eliminates the hub effect and homophily, the number of cases decreases by 11.4%. This result indicates that the homophily effect appears to be the dominant factor in this scenario.

From the right upper panel, the conclusions on the advantages of our proposed strategy also remain robust. Our proposed approach still reduces the cases by three times more, compared to the random targeting or uniform targeting strategies. It also outperforms the strategy that targets the most central CBGs. These results are consistent with those of January 21st, 2022, though here the effect sizes are generally larger for July 1st, 2021 as the vaccine was assumed with perfect efficacy.

From the lower panel, we find the targeted CBGs that are largely consistent with that in the main text. In particular, among all CBGs with a targeted increase in the vaccination rate by 5%, the Jaccard similarity is as much as 76.5%; among the targeted CBGs on January 21st, 2022, 87.0% were targeted on July 1st, 2021. These results continue to show our robustness of our proposed targeting strategy under different spatial vaccine distributions, variants, or vaccine efficacy levels.

### Illustrations for synthetic networks

We first provide a simple illustration for our synthetic networks (of CBGs). Note that we first construct bipartite networks between CBGs and POIs, and next convert them to networks of CBGs. To simplify visualization, We present networks of 50 nodes only, although our simulation utilizes 10,000 nodes. The network of CBGs are very dense, so we filtered out the links with the smallest weights for the visualization purpose. The left panel of Fig. S2 presents a centralized network where well-connected hubs tend to have higher vaccination rates. The middle panel illustrates a network of two clusters – one with high and another with low vaccination rates. The right panel presents a network with a mix of edges from both the centralized and the clustered networks. This network has two clusters with different overall vaccination rates. At the same time, it also exhibits centralization with higher vaccination at hubs of each cluster than other non-central nodes of the same cluster.

**Figure S1.**
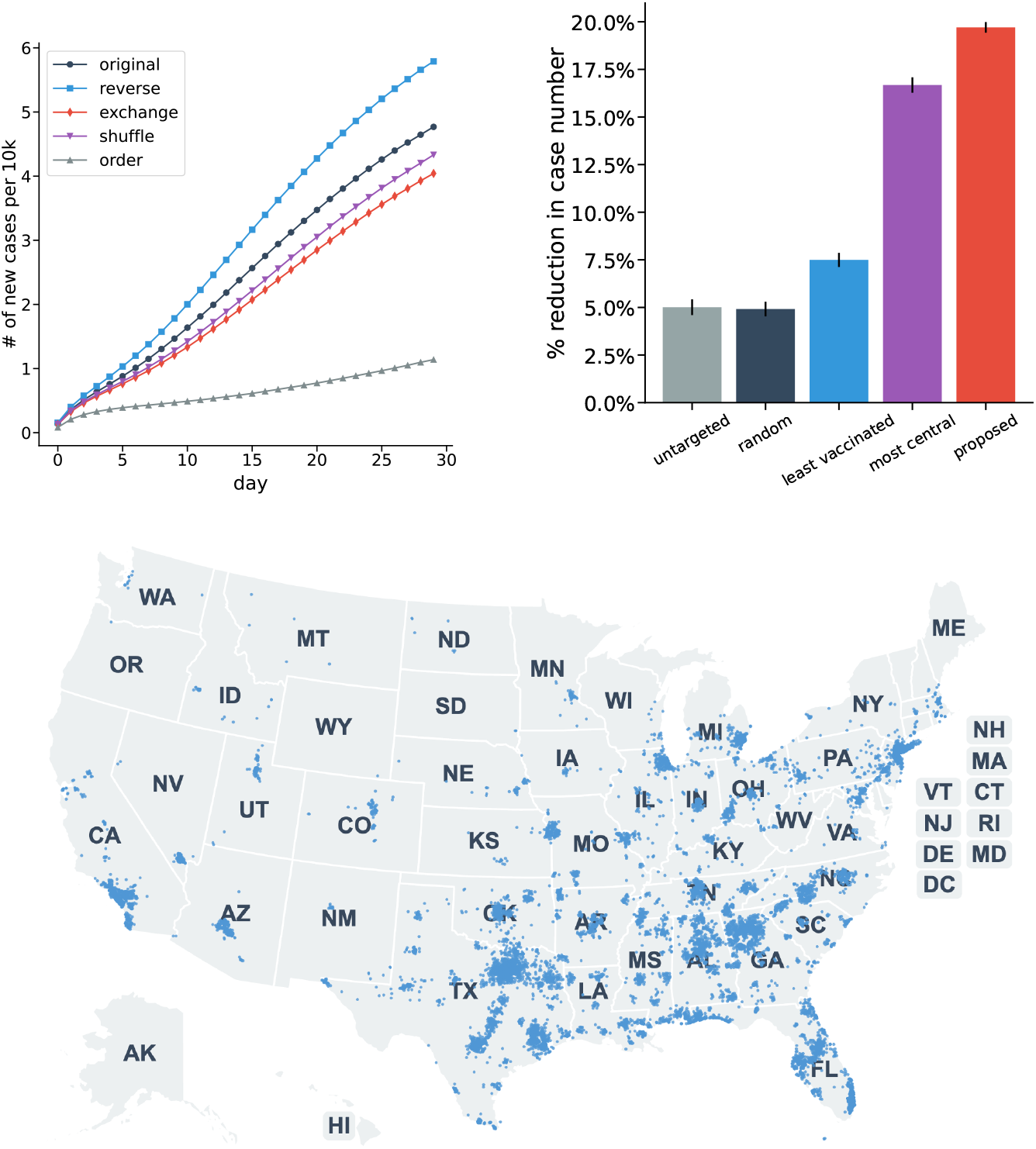
The replication result for July 1st, 2021, without consideration of reinfection, breakthrough infection, or imperfect vaccine. (**Upper left**) The number of simulated cases based on the U.S mobility network under multiple hypothetical scenarios. Error bars are standard deviations of simulations. (**Upper right**) The performance of the four targeting strategies. The *y*-axis is the reduction compared to the current vaccination distribution (“original”) in the first 30 days. Error bars are standard deviations from different runs of simulations. (**Lower**) The most strongly targeted CBGs whose vaccination rate increases more than 5% under a scenario with our proposed strategy. Each blue dot represents a targeted CBG with more than 5% targeted increases.

**Figure S2.**
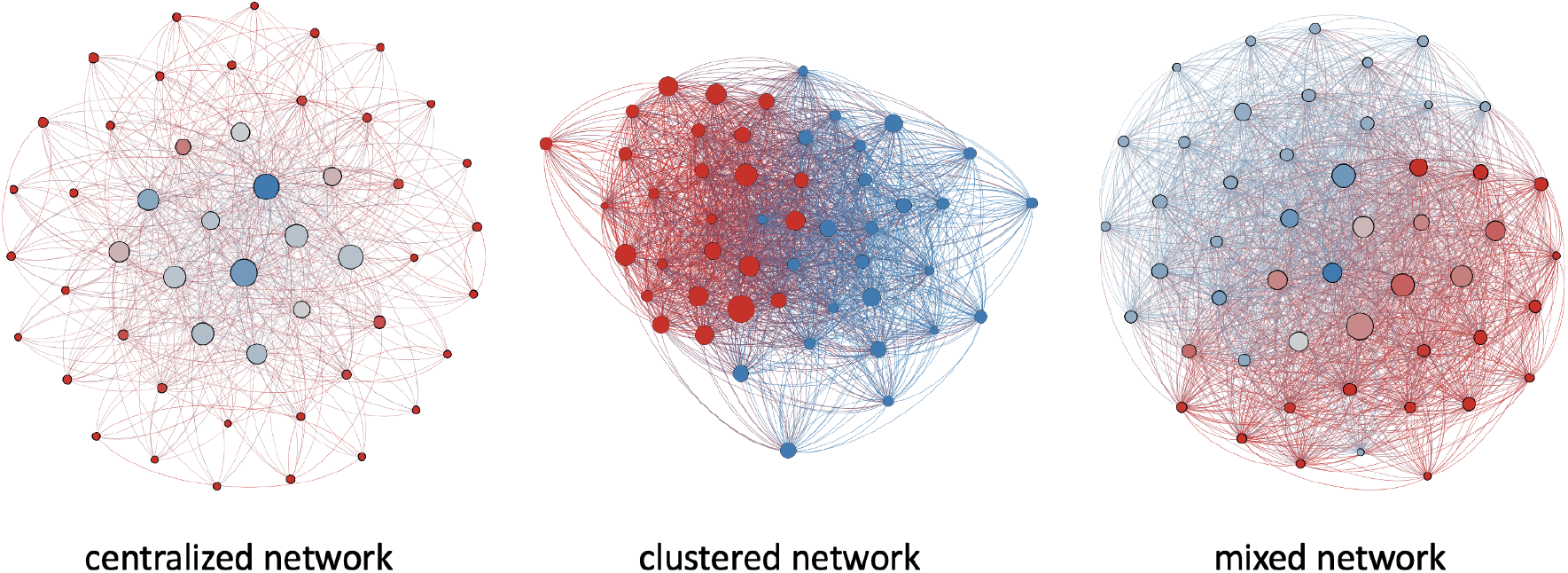
Simple illustrations of the synthetic centralized network, clustered network, and the mixed network (with *λ* = 0.5) of CBGs. Blue and red indicate high and or low vaccination rates, respectively. Larger nodes have higher mobility centrality in the network. Nodes are positioned according to the Fruchterman-Reingold layout^37^.

### Details for the parameters in the simulations

Our simulations are based on ref. 24 with modifications to adapt to the goals of our study. For the synthetic networks, we run the simulations over 30 days by setting the initial infection rate to 0.01% and the transmission rate to 0.1. The average natural immunity period and vaccine wear off period are set as 90 days. These parameters have no specific meanings and their choice would not change our main conclusions other than the growth rates in Fig. 2. The simulations on the synthetic networks enforce a within-CBG transmission rate of 0 since our focus is on how cross-CBG transmissions affect the eventual case number.

For the U.S. country-level simulation, we set the initial infection rate to 0.1%, the country-wide cross-CBG transiting to *ϕ* = 1500 (multiplied by a POI’s factor) and within-CBG transmission to 0.005 (these numbers affect the transmission rates). The average natural immunity period and vaccine wear off period (*l*_imm_) are set as 90 days for the state of January 2022; The vaccine efficacy (*α*) is set to be 0.7. The choice of these values is informed by their estimates in the ten major metro areas studied in ref 24. Marginal changes to these values would not alter our main conclusions significantly. To account for different transmission dynamics, *l*_imm_ = ∞ and *α* = 1 when we simulate the state of July, 2021.

Ref. 24 employs inferred hourly mobility patterns to conduct simulations which aim to most accurately predict the growth of COVID-19 transmission. By contrast, our study aims to examine how the homophily and hub effects of heterogeneity in vaccination affect the frequency of infections when human mobility returns to the pre-pandemic levels. Thus, the input to our simulations is the hourly average number of visits in 2019 rather than their inferred values above. All our results, including Figure 3, are based on the simulations over a period of 30 days.

**Figure S3.**
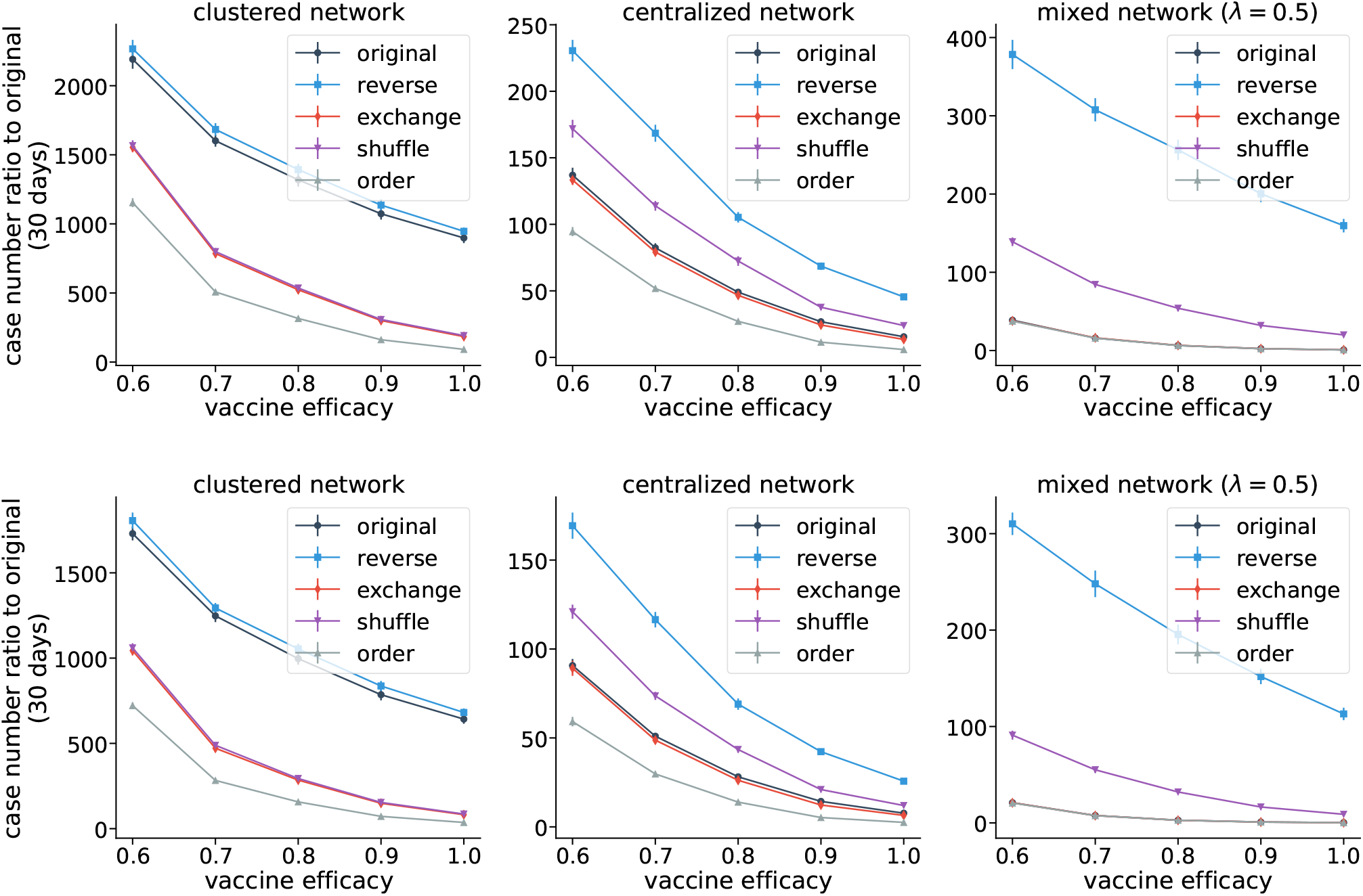
**(Upper)** The simulation for different hypothetical distributions for different networks *with* accounting for reinfection/breakthrough infection scenario. **(Lower)** The simulation for different hypothetical distributions for different networks *without* accounting for reinfection/breakthrough infection scenario. The *x*-label is the vaccine efficacy. Each curve represents hypothetical distributions under different levels of vaccine efficacy.

### Robustness of simulation results

Figure S3 presents the results for five hypothetical distributions under different levels of vaccine efficacy. The upper and lower panels represent the scenario with or without reinfection/breakthrough infection scenario, respectively. These results show that our main conclusions are consistently robust — regardless of vaccine efficacy, or the consideration of reinfection and breakthrough infection, the relative magnitudes of the five hypothetical distributions are consistent. These simulation results also suggest that in real world scenarios our conclusions on the two network effects would be also likely robust to different transmission dynamics variants and vaccine efficacy levels.

### Mobility centrality

In network science, there are multiple measures for node centrality^52^. They describe the degree to which each node is central in the network under different contexts. In our study, we employ the weighted degree centrality. Specifically, we use *W* to denote the weighted adjacency matrix (|*𝒞* | ×|*𝒞* |). Then, mobility centrality (MC) is defined as

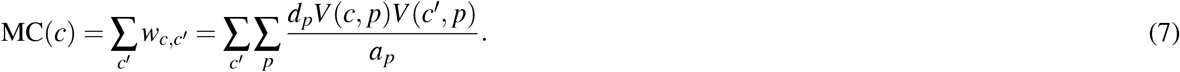

Intuitively, a more mobile and populous CBG, or a CBG connected to many other CBGs (through mutually visited POIs), should have a higher mobility centrality score. There are different ways of defining the edge weights. We choose this edge weight because it directly reflects the extent of transmission between two CBGs, as it corresponds to Eq. (2). Thus, a more mobile CBG is considered more central as it is more vulnerable to contracting the disease. Similarly, there are other valid choices for the centrality score^52^. However, since our study examines a mobility network of more than 200, 000 CBGs, calculating other centrality measures (such as eigenvector centrality or betweenness centrality) becomes computationally expensive. Nevertheless, as previous work has shown, degree centrality is highly correlated with other centrality measures, specifically eigenvector centrality^53^. Thus we do not expect the choice of centrality measure to significantly change our conclusions.

Figure S4 presents the mobility centrality maps of California and Texas, the two most populous states in the U.S. As shown in the figure, CBGs that are closer to the large cities (such as Los Angeles and San Francisco in California and Dallas and Houston in Texas) have larger centrality scores. Moreover, CBGs in Texas have larger average mobility scores than those in California, indicating that residents in Texas are on average more mobile than in California.

### Measuring homophily in U.S. data

Here, we analyze the correlation between a CBG’s vaccination rate and the weighted average among its neighboring CBGs in the U.S. This analysis is complementary to the simulations discussed in the main text and provides direct evidence on the existence of homophily in the US, yet it does not capture the impact of homophily on the number of infections. Fig. S5 provides an intuitive visualization of the strength of the homophily effect. As shown in the Figure, there is a strong positive correlation between a CBG’s vaccination rate and its neighbors’ weighted average (*ρ* = 0.845, *p <* 0.001). This strong positive correlation suggests a high level of clustering by vaccination, dense clusters of high and low vaccination CBGs with many connections within and few connections between, which may lead to more infections compared to a uniform vaccine distribution without the homophily effect. Figure S5 also shows this correlation under the “exchange” distribution. The correlation is largely reduced, thus confirming the rationale behind the “exchange” procedure which largely reduces the homophily effect.

### Measuring hub effect in U.S. data

Similar to the previous section, here we present results of a complementary analysis to the simulations which verify the existence of the hub effect. We first divide all CBGs by their mobility centrality^52^ into deciles, and then plot the average vaccination rate in each decile. Furthermore, to understand the within-state relationship between mobility centrality and the vaccine rate, we plot the vaccination of CBGs versus their within-state centrality *z*-scores. The country-level and within-state binned scatter plots are presented in Fig. S6.

**Figure S4.**
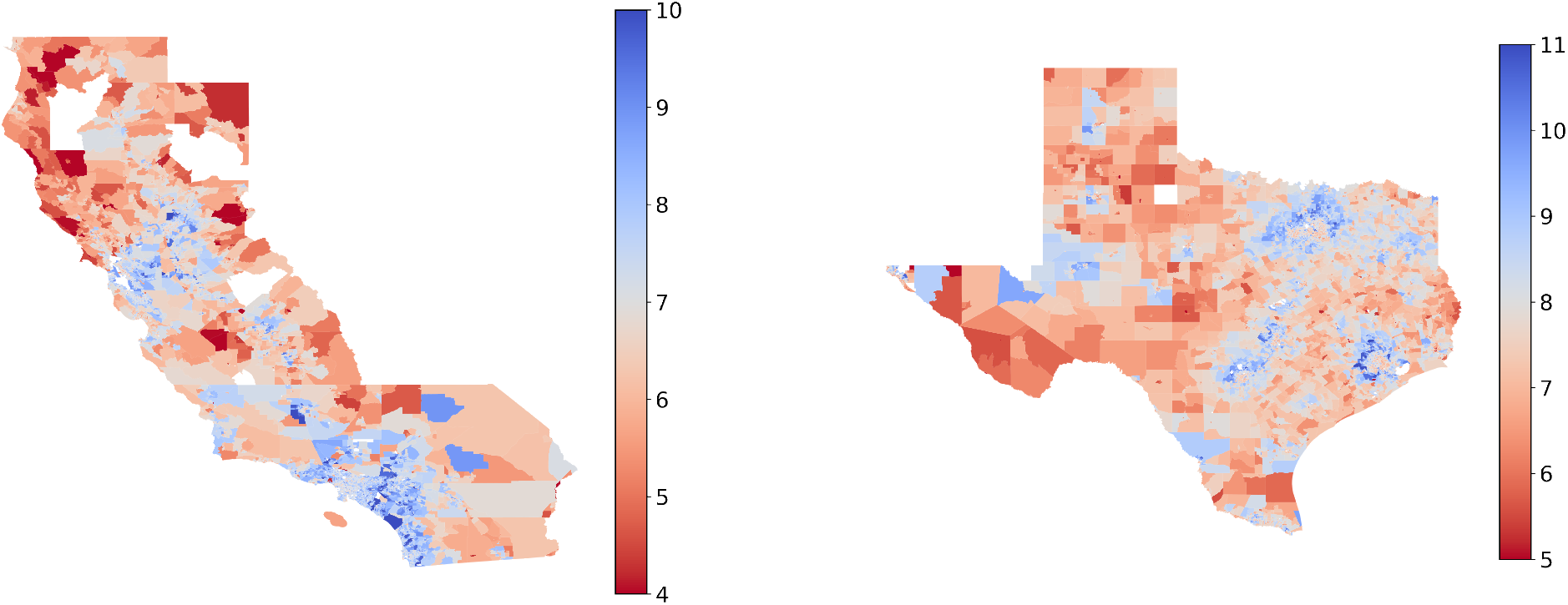
The mobility centrality maps for the two most populous states in the U.S. The centrality is presented and color coded in log-scale. We present the California and Texas as examples. In general, urban and populous areas tend to be more central.

**Figure S5.**
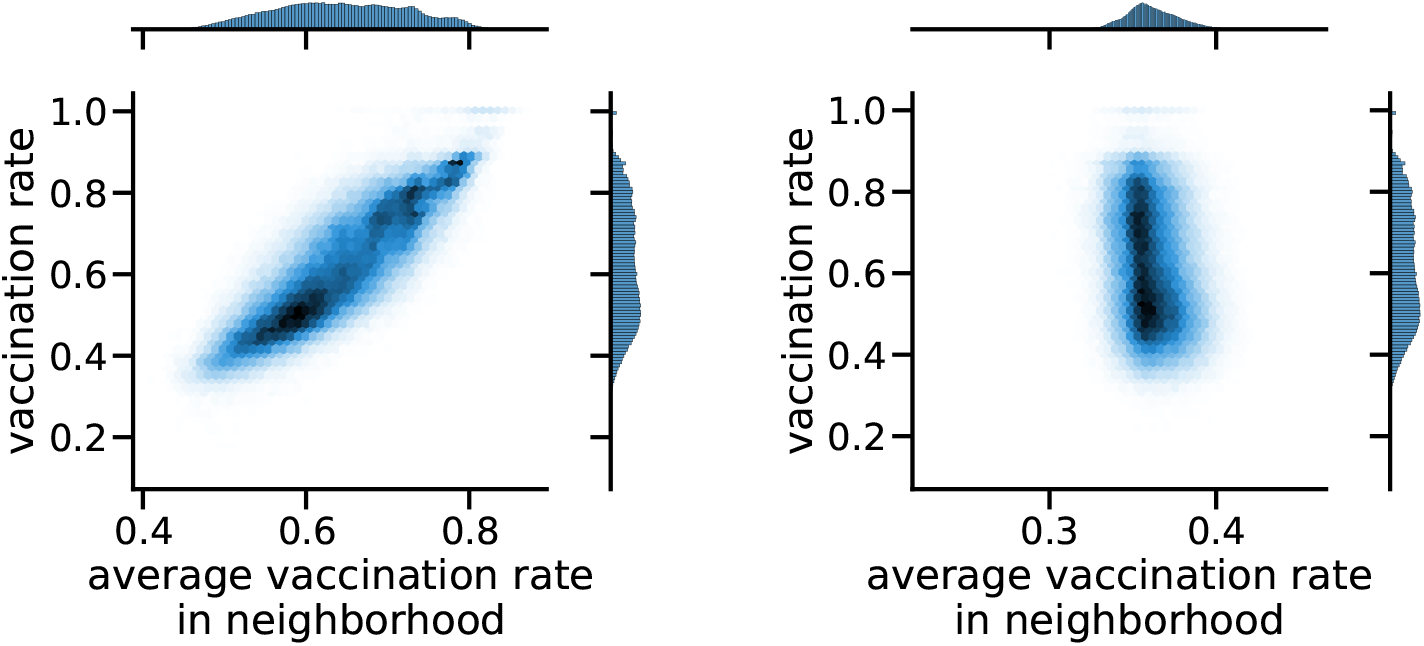
**Left**: The vaccination rate of a CBG versus the average among its neighbors in the network weighted by their mobility centrality under the <u>“original”</u> distribution. **Right**: The same plot as the one on the left, but under the <u>“exchange” distribution</u>.

In the left panel, we see that as centrality in the country network increases by one decile, the average vaccination rate increases by 1.0%. This result suggests a positive correlation between centrality and vaccination. In the right panel, we observe that as within-state centrality increases by one decile, the average vaccination rate increases by 1.2%. Such a “reverse” distribution also reflects the urban-rural divide in vaccination that is common in many states.

**Figure S6.**
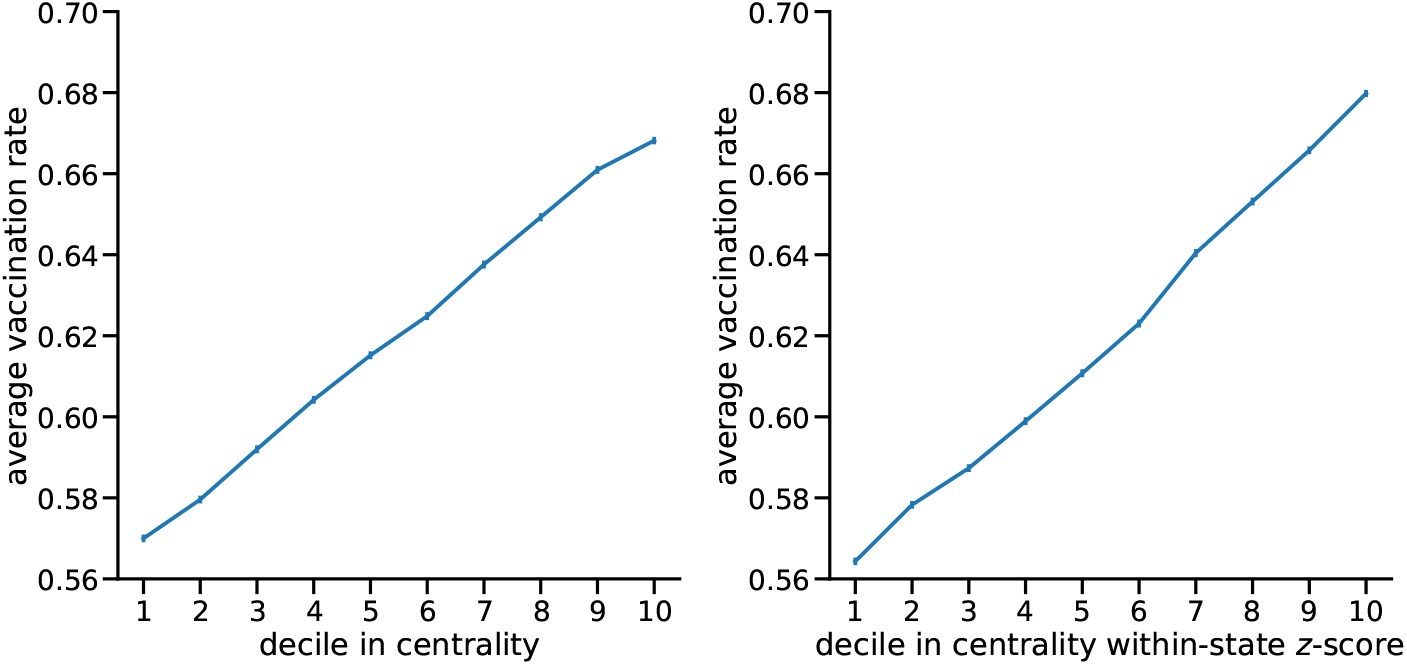
The average vaccination rates in each decile of the country-wide mobility centrality (left) or the normalized centrality *z*-score within states (right). Error bars are standard errors of the averages.

### Within-state hypothetical vaccination distributions and simulations

The hypothetical simulation analysis in this section is similar to the one presented in the main text, with the exception that the hypothetical distribution generating procedure is conducted within each state separately, while removing any cross-state mobility from the network. Thus we perform the simulation separately in each of the fifty state and district networks (except Hawaii for which we don’t have vaccination data). The state networks exhibit large variations in their structural properties, which may imply the generality of our results to other regions, especially less populous countries. The results in this section can thus provide extra insights on how spatial heterogeneity in vaccination, especially the homophily and hub effects, affects the overall transmission in different regions.

As shown in the upper left panel of Fig. S7, we find that many states exhibit a significant increase in the case count by reversing the vaccination rates, with Kansas, Nebraska, and Indiana being the top three states. As for the homophily effect, we find different patterns than the simulation result for the whole country. That is, for many states, the “exchange” distribution does not significantly reduce the cases. We conjecture that this may be due to the following reasons. First, the “exchange” procedure may only succeed in removing the homophily effect in a large region (e.g. the entire U.S.). If we exchange the vaccination rates of CBGs with similar centrality scores in a small region, we may not sufficiently shuffle the vaccination rate such that homophily is largely removed. These results imply the limitations of our exchange approach as it potentially retains the homophily effect if applied to small regions. This echoes our results in the synthetic networks: if the hub effect dominates, the exchange distribution may not clearly show the homophily effect.

Combining with “shuffle”, we also obtain insights into the effects of the homophily effect. For each state and each hypothetical distribution, we run 25 rounds of simulations and obtain the case counts. We then generate the “reverse:original” ratio, “exchange:original” ratio, and “shuffle:original” ratio. We treat “shuffle:original” ratio as the dependent variable and “reverse:original” ratio and “exchange:original” as independent variables. With an ordinary least squares model, we find the regression coefficient for the “reverse:original” is 0.343 (*p <* 0.001) and the regression coefficient for the “exchange:original” is 0.377 (*p <* 0.001). This result indicates that both the hub effect (“reverse”) and the homophily effect (“exchange”) contribute to the shuffled result. Since “reverse:original” ratios are generally larger than the “exchange:original” ratios, the “shuffle” result (for example the ranks) is consistent with the “reverse” result in the main text.

From the lower right panel, we see that although the current hub effects are shown to be strong from the “reverse” effect, we can still leverage and increase the hub effect to further reduce the case number. For example, we find that by improving the hub effects in states such as Arizona, Texas, and Louisiana, we would observe very large reductions in the case counts.

### Details of inferred CBG-level vaccination rates

Deep neural networks (deep learning) have been shown to be powerful in predicting fine-grained level statistics, such as poverty^54^. Here we discuss more details about our Bayesian neural network. We use a three-layer network with ReLU activation. We then assume a Gaussian prior on the parameters of the neural network Θ^35^. Exact inference over the posterior of a Bayesian neural network is intractable, so we use an approximate inference technique based on dropout^36^. Moreover, we run multiple inferences with random initialization and take the average of resulting vaccination rates for each CBG, which is referred to as the “neural network ensemble” model.

Fig. S8 presents our inferred results of the two most populous states: California and Texas. The left panels of each row present the inferred CBG level vaccination rate. To illustrate how the inferred vaccination rate reflects demographic features, we plot the estimated average age and education level (percentage with Bachelor’s or higher degree) for each CBG as examples. As shown in Figure, the CBGs with higher vaccination rates in general have either a higher average education level or higher average age.

As mentioned in the *Methods and Materials* section, although we do not have the CBG-level groundtruth of vaccination rates to validate our predictions, a few states of the U.S. released the zip-code level vaccination rates. These states include California, Idaho, Illinois, Maine, New York, Oregon, Pennsylvania, and Texas as of January 21st, 2022. We thus resort to use the zip code level vaccination rates to show the superiority of our approach compared with baseline models.

We compare with three baseline methods in Figure S9: **mean**: we use the (directly available) county-level vaccination rate to impute the CBG-level vaccination rate; **linear**: we use linear regression as the mapping from features to vaccination rates; **logistic**: we use logistic regression as the mapping from features to vaccination rates. A single network network is labeled as “NN” and the average of multiple neural networks’ inferences is labeled as “ensemble”. We measure the mean absolute error (MAE) weighted by population to evaluate the performance. From this figure, we observe that our neural network based method achieves lower weighted MAE than all of the baselines, and ensembling multiple randomly initialized neural networks can further improve performance. Therefore, we use the ensemble version as the inferred CBG-level vaccination rates in the main text.

**Figure S7.**
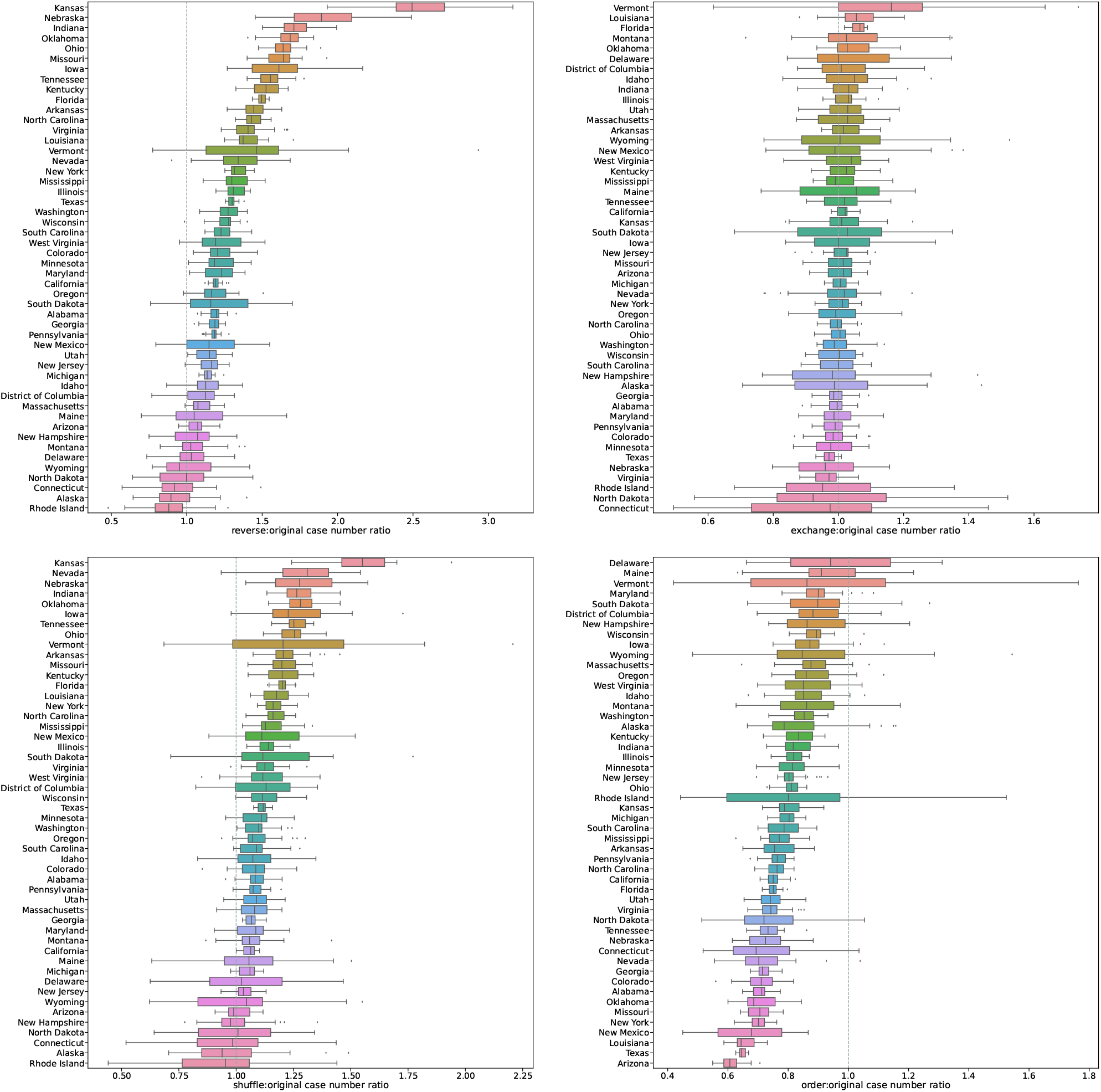
The box plots of the ratio of (**upper left**) “reverse” to “original”, (**upper right**) the ratio of “exchange” to “original”, (**lower left**) the ratio of “shuffle” to “original”, and (**lower right**) the ratio of “order” to “original” distributions within each state. States are ranked by the ratio averages. In boxplots, center line, box limits, whiskers, and points represent medians, upper or lower quartiles, 1.5x interquartile ranges, and outliers, respectively.

**Figure S8.**
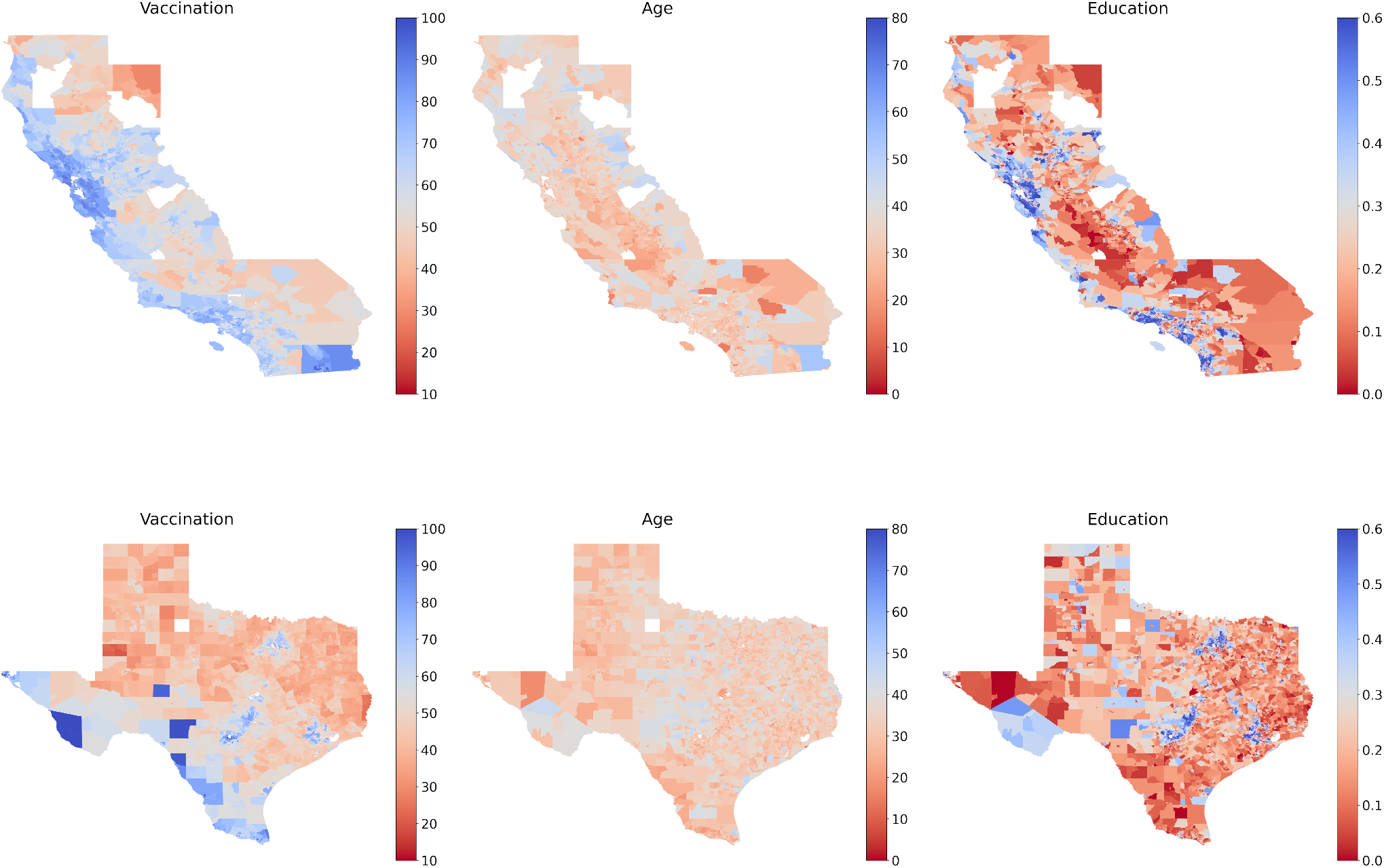
Inferred vaccination rates on the two most populous states. Blue indicates higher vaccination rate (or higher average age or education level) and red indicates low vaccination rate (or higher average age or education level). For comparison, we also plot the average age and the fraction with college degree. In general, CBGs with either a higher average age or a higher education level have higher vaccination rates. This is consistent with the observation that in general older people have higher vaccination rate (because of earlier access) and better educated people have higher vaccination rate.

### Details of the proposed vaccination strategy

We solve the optimization problem by projected gradient descent^41,42^ At each step, we take a gradient step to minimize (*u* - *v*)^*T*^*W* (*u* - *v*). The resulting *v* might be infeasible, i.e. fail to satisfy the constraints in Eq.(5,6), so we project *v* back to the feasible set. In particular, to satisfy Eq.(5) we can compute the projection by

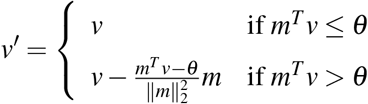

To satisfy Eq. (6) we can compute the projection by

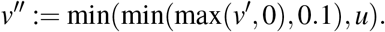

Intuitively, we lower bound *v*_*c*_ by 0 and upper bound it by the smaller of 0.1 and *u*_*c*_.

Formally, the algorithm is as follows:

1. Initialize *v*^0^, *λ* ^0^ = 0, *γ*^0^ = 0;
2. For *t* = 0, …, *T* :
  a. *v*^*t*+1^ := *v*^*t*^ + *η*_*t*_(2*W* (*u* - *v*^(*t*)^);
  b. Set *v*^*t*+1^ := min(min(max(*v*^*t*+1^, 0), 0.1), *u*);
  c. Set 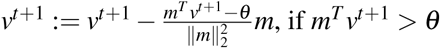.

**Figure S9.**
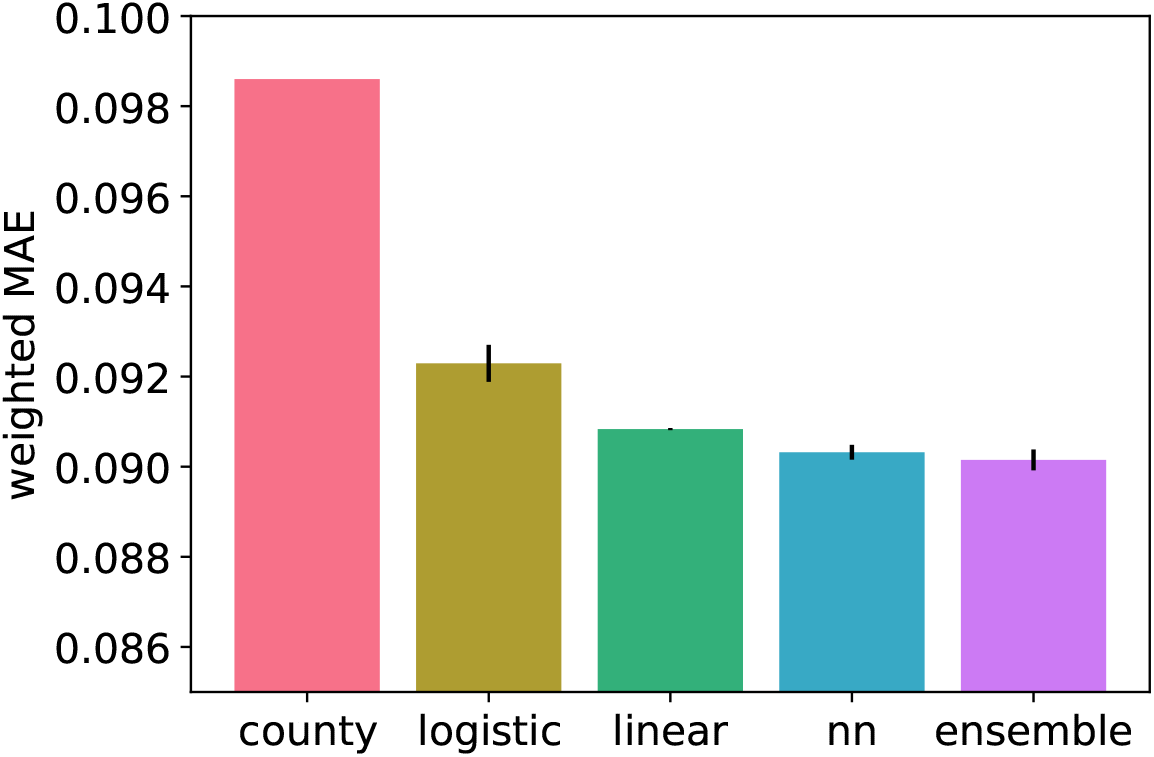
The mean absolute error (MAE) weighed by population on the zip code level vaccination rate. Small area estimation (linear, logistic, neural networks and ensemble) achieves smaller MAE compared to directly using county-level data (county). Among the estimation methods, neural network (NN) achieves the smaller MAE compared to linear regression (linear) or logistic regression (logistic), and ensembling multiple neural networks further improves performance.

The algorithm must converge with a suitably selected learning rate *η*_*t*_ based on standard results in optimization theory^41,42^ (i.e. because each step in the algorithm does not increase the L2 distance to the optimal solution). Upon convergence, the resulting *v*_*T*_ is the optimal solution (*v*^*^) to the optimization problem in Eq. (4), as shown by the following theorem.

**Theorem 1**. *If we choose* 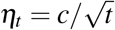 *for any c* ∈ ℝ^+^, *the algorithm above converges to the global optimum of the optimization problem in Eq.(4)*.

*Proof of Theorem 1*. We first prove that the optimization problem is convex. First, observe that the matrix *W* in Eq.(4) is a positive semi-definite matrix. This is because there exists matrix *U* such that *W* = *UU*^*T*^. Concretely, we can construct *U* by

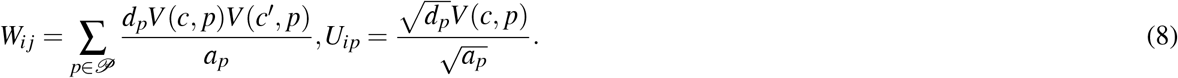

Second, Eq.(5) is a linear inequality, and Eq.(6) are both linear inequalities. Therefore, the objective Eq.(4) and the constraints Eq.(5, 6) are all convex or linear, hence the problem is convex.

In adition, because the optimization objective Eq.(4) is a Lipschitz function, therefore, by standard results^55^ projected gradient descent converges to the global minimum of the optimization problem. □

Finally, we plot the histogram of the vaccination rate increases for all the CBGs in Fig. S10. As shown in the figure, our proposed strategy targets the approximately same number of CBGs with the random targeting strategy or the lowest-vaccination-rate-CBG targeting strategy. Much fewer CBGs are targeted but have an increase smaller than 10%. Targeting the most central CBGs involves targeting a slightly fewer number of CBGs because the average sizes of these most central CBGs are in general larger.

**Figure S10.**
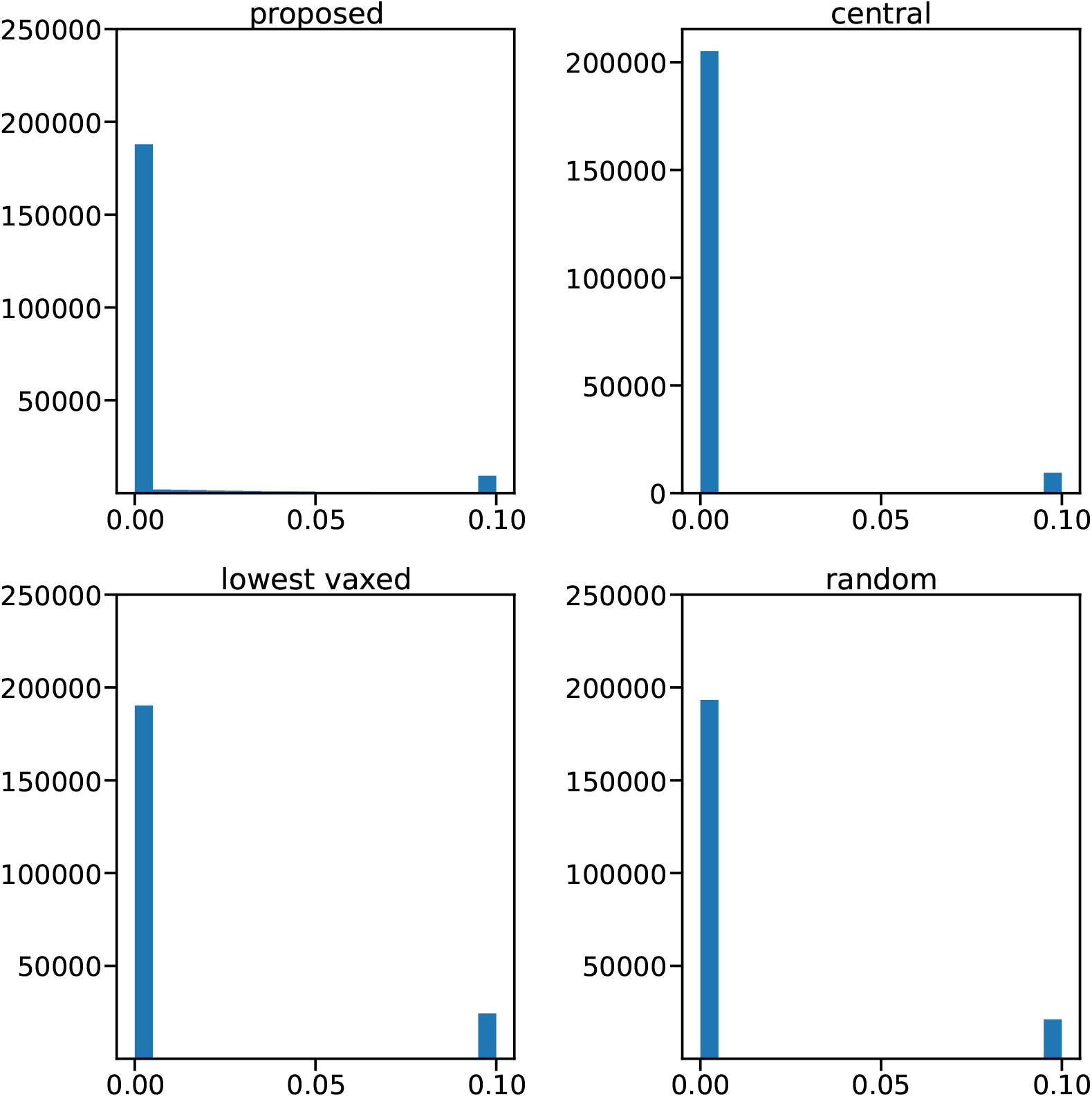
Histogram of the vaccination rate increases for all the CBGs (0 if untargeted). The histogram is a bi-modal distribution for all strategies (except for uniform, which is 1% for all CBGs): for the majority of CBGs the vaccination rate increase is 0% (i.e. these CBGs are not included in the vaccination campaign), while for a small proportion of CBGs the vaccination rate increase is 10% (which is the maximum vaccination rate increase that we assume is feasible).

## Footnotes

a For the later U.S. simulations, we apply the reverse distribution to each state separately, which prevents conflating variation in the mobility centrality of different states and helps retain the urban-rural divide in vaccination within states. Also note that due to large differences in vaccination rates across states, conducting the reverse distribution across the country may simply capture the effect of heterogeneity in vaccine acceptance among the states with high and low rates, which is not the main point of this paper.

b In practice, we exchange the first and the second, the third and the fourth, etc.

c Here, we assume that the cost of promoting additional 1% to get vaccinated is the same across locations, which is likely not the case. Given the lack of theories on this, we make this simplification for the sake of the simplicity.

d https://covid.cdc.gov/covid-data-tracker

e The vaccination data from Hawaii is not available and Hawaii is not included in our analysis. Given that their population makes up a tiny fraction and that Hawaii is an island state, we believe that its impact on the country-level outcomes could be marginal or negligible.

f In practice, each CBG has an index from 1 to 10,000 which is independent of its own attributes, and we match on the index.

g We acknowledge political ideology is also predictive, but we cannot use them to impute CBG-level vaccination rates as turnout data are not available on the CBG level.

